# Predictive accuracy of diagnostic tests for excessive bleeding in cardiac surgery: the COPTIC-C study

**DOI:** 10.1101/2024.10.17.24315651

**Authors:** Weiqi Liao, Robert Grant, Florence Y Lai, Hardeep Aujla, Marcin Wozniak, Hasmukh R Patel, Laura Green, Andrew Mumford, Gavin J Murphy

## Abstract

**Purpose:** We tested the hypothesis that addition of biomarkers of multimorbidity and biological ageing would improve the predictive accuracy of point-of-care viscoelastometry or laboratory tests of coagulation for clinically important bleeding following cardiac surgery.

**Methods:** The analyses included 2437 participants in the Coagulation and Platelet laboratory Testing in Cardiac surgery (COPTIC study) with complete clinical, TEG®5000 Thromboelastography, ROTEM®, Multiplate® aggregometry, full blood count, laboratory reference tests of coagulopathy, and biomarkers of biological ageing and multimorbidity. Models with different biomarkers to predict the composite primary outcome, Clinically Important Bleeding, was developed using logistic regression and internally validated using 10-fold cross-validation. Discrimination, calibration, and clinical utility of the models were assessed comprehensively.

**Results:** For the primary outcome, the AUROC for the best predictive model using TEG/ROTEM with other biomarkers was 0.694 (0.612–0.775). The best predictive model included laboratory reference tests of coagulation, full blood count results, and biomarkers of multimorbidity and ageing, AUROC=0.701 (0.620–0.781), although clinical utility was not superior to using laboratory reference tests alone. Discrimination was higher for components of the primary outcome; large volume (≥4 units) red cell transfusion 0.754 (0.602–0.903), and large volume procoagulant transfusion 0.723 (0.590–0.857), but not for excess loss in drains/re-sternotomy 0.701 (0.613–0.788). Calibration was generally good among the models.

**Conclusion:** Diagnostic tests for bleeding following cardiac surgery demonstrate moderate discrimination, although this was influenced by the definition of bleeding. Small improvements in discrimination with inclusion of additional disease biomarkers, with similar calibration and clinical utility.

**Take-home message:**

1. Current diagnostic tests demonstrate moderate predictive accuracy for excessive bleeding following cardiac surgery. In this study, addition of biomarkers of multimorbidity and biological ageing improved discrimination but not clinical utility.
2. Existing clinical definitions of bleeding represent heterogeneous phenotypes, presenting a barrier to research investigating the disease processes.

Important abbreviations used in this paper:

- CIB – Clinically Important Bleeding
- CCB – Clinical Concern about Bleeding
- AUROC – Area Under the Receiver Operating Characteristic Curve

## Introduction

Excessive bleeding is common following cardiac surgery where it increases the risks of postoperative infection, organ injury, and death. [1, 2] Clinical interventions targeting excessive bleeding do not improve clinical outcomes beyond reductions in bleeding and transfusion requirement. [3, 4] The imprecision of existing clinical definitions of bleeding and diagnostic tests are barriers to research addressing this paradox.

Consensus definitions of bleeding focus on clinical parameters such as volume of transfusion or blood loss, or requirement for reintervention [1, 2, 5–7] rather than disease mechanisms, which are poorly understood. In the COagulation and Platelet laboratory Testing in Cardiac surgery (COPTIC) study (ISRCTN20778544), and a subsequent meta-analysis of COPTIC and similar studies, we demonstrated that laboratory reference tests of coagulation were poor predictors of bleeding, as were point-of-care tests of coagulopathy based on viscoelastic tests (i.e. thromboelastography [TEG] and rotational thromboelastometry [ROTEM]), or platelet aggregation. [6] These observations led us to hypothesise that alternative disease mechanisms that cause bleeding were unmeasured in these analyses. This was supported by a secondary analysis of COPTIC data where biomarkers of multimorbidity and biological ageing were significantly associated with the study primary outcome, Clinical Concern about Bleeding (CCB). [8]

To improve our understanding of bleeding phenotypes, this study sought to test the hypothesis that the predictive accuracy of TEG/ROTEM or laboratory reference tests of coagulopathy would be improved by the addition of baseline biomarkers of multimorbidity or biological ageing. A secondary hypothesis evaluated whether the clinical definition of the bleeding outcome influenced the results.

## Methods

### Study design and data source

The predictive accuracy study design minimised the risk of bias for the domains identified in QUADAS-2 ^1^. The protocol was prospectively registered (ISRCTN20630689)^2^, and was reported following the STARD guidelines. [9] The development of the predictive models was reported following the TRIPOD statement [10] (both checklists in the appendix). The UK Human Research Authority ethics approved this study (Ref: 21/NW/0157).

Patients recruited to the COPTIC study between March 2010 and September 2012 who provided written informed consent for secondary analyses were eligible. The COPTIC clinical, TEG, ROTEM, Multiplate, Sysmex and laboratory tests of coagulopathy datasets were linked to biological ageing and organ dysfunction biomarkers by unique participant ID. 38 Participants without available lab results were excluded (**eTable 1**).

### Index tests

The test data included **1.** Post Protamine Point-of-care TEG (TEG® 5000 Thromboelastograph®, Haemonetics, Coventry, UK) and INTEM, EXTEM, FIBTEM, and HEPTEM tests using ROTEM® (Werfen, Cheshire, UK) in whole blood, **2.** Post Protamine laboratory tests of coagulation; PT, APTT, Clauss Fibrinogen activity, vWF Ristocetin cofactor activity, FXIII activity, D-dimer concentration, anti-Xa assay using an unfractionated heparin standard, and thrombin generation. **3.** Post Protamine platelet aggregometry in whole blood (ADP-test®, ASPI-test®, TRAP-test®, Adren-test®) used the Multiplate® device, and **4.** Post protamine full blood count (FBC) was measured using the Sysmex X2100 Analyser, all performed at the time of the primary study, plus **5.** Pre-surgery biomarkers of multimorbidity (organ dysfunction); serum bilirubin, alkaline phosphatase, full blood count, glomerular filtration rate (eGFR) estimated from serum creatinine, transferrin, ferritin, interleukins (IL)-6, IL-8, high sensitivity serum Troponin I and pro-NT BNP and **6.** Biomarkers of biological ageing including CCL11/Eotaxin, CX3CL1/Fractalkine, GDF-15, IL5, and CXCL9/iAge, all measured in thawed from frozen, double spun plasma collected during the primary study and stored at −80⁰C until batch analyses. The methods for these assays have been described in detail previously. [6, 8]

### Clinical outcomes

The pre-specified primary outcome was Clinically Important Bleeding (CIB), defined as ANY of the following events:

1. **Large volume red cell transfusion** (≥4 units) from the administration of protamine to 24 hours post-surgery;
2. **Large volume transfusion of pro-coagulants** or clotting factors; FFP transfusion >2 units, cryoprecipitate transfusion >2 units, platelet transfusion >1 unit, or use of recombinant Factor VII from the administration of protamine to 24 hours post-surgery.
3. **Severe blood loss** was defined as loss in drains >600ml in 4 hours post return to ICU, or emergency re-sternotomy for bleeding, where a ‘surgical’ cause had been excluded.

### The secondary outcomes included

a. Individual components of the primary outcome (above),
b. Postoperative Clinical Concern About Bleeding (Postop CCB) as defined in the original COPTIC study, [6]
c. A composite clinical outcome of in-hospital death, KDIGO defined Stage 2 or Stage 3 acute kidney injury, myocardial infarction, stroke, and infection, occurring within the index hospitalisation, all as defined in the primary publication. [6]

### Participant flow and clinical management

A schematic of participant flow through the study is shown in **Figure 1**. Two 22.5 ml blood samples (45ml in total) were obtained in the operating theatre for research at the following time points:

i. Immediately before induction of anaesthesia (pre-surgery),
ii. Twenty minutes post-reversal of heparin anticoagulation (post-protamine, before chest closure).

**Figure 1.**
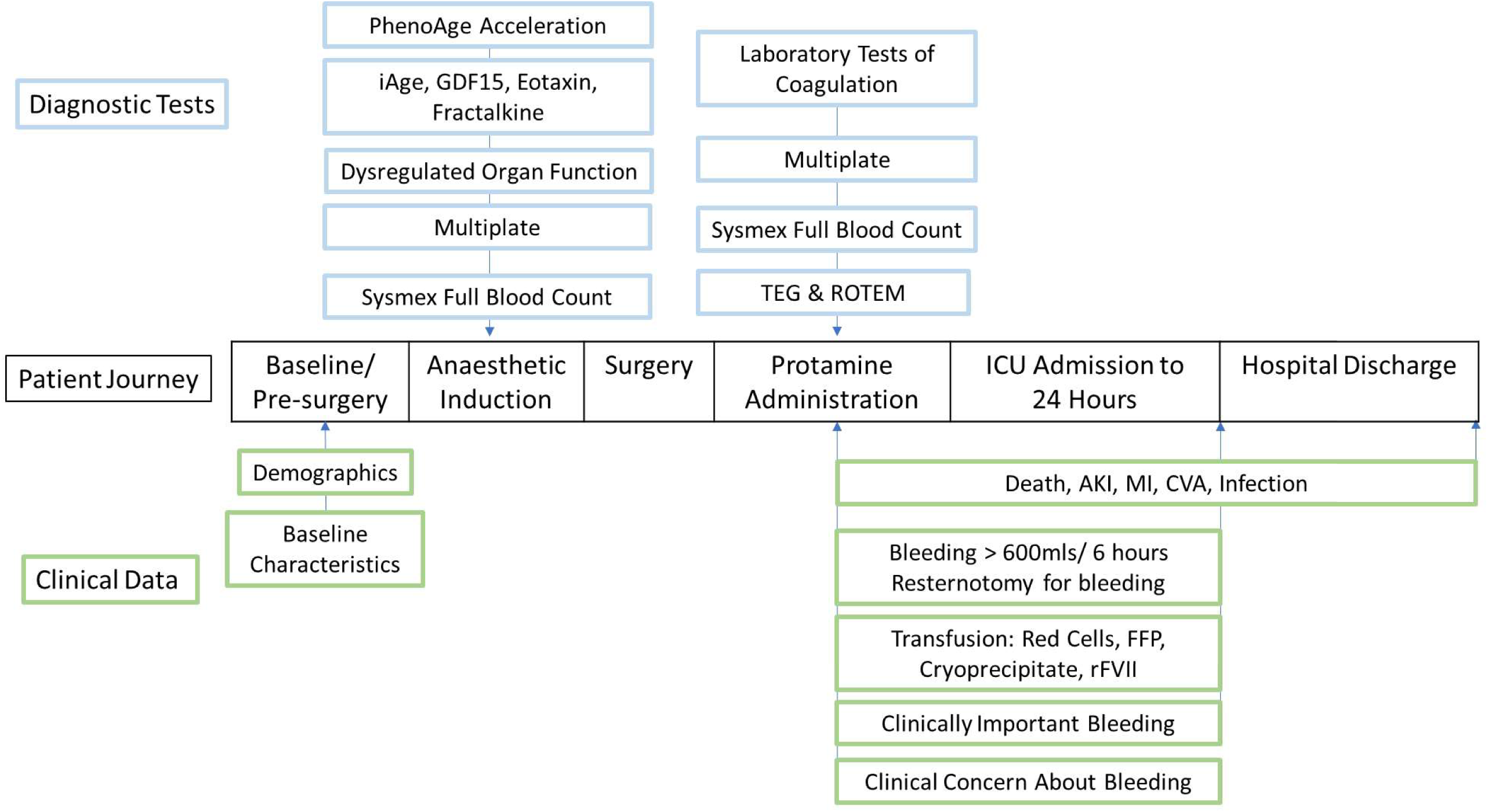
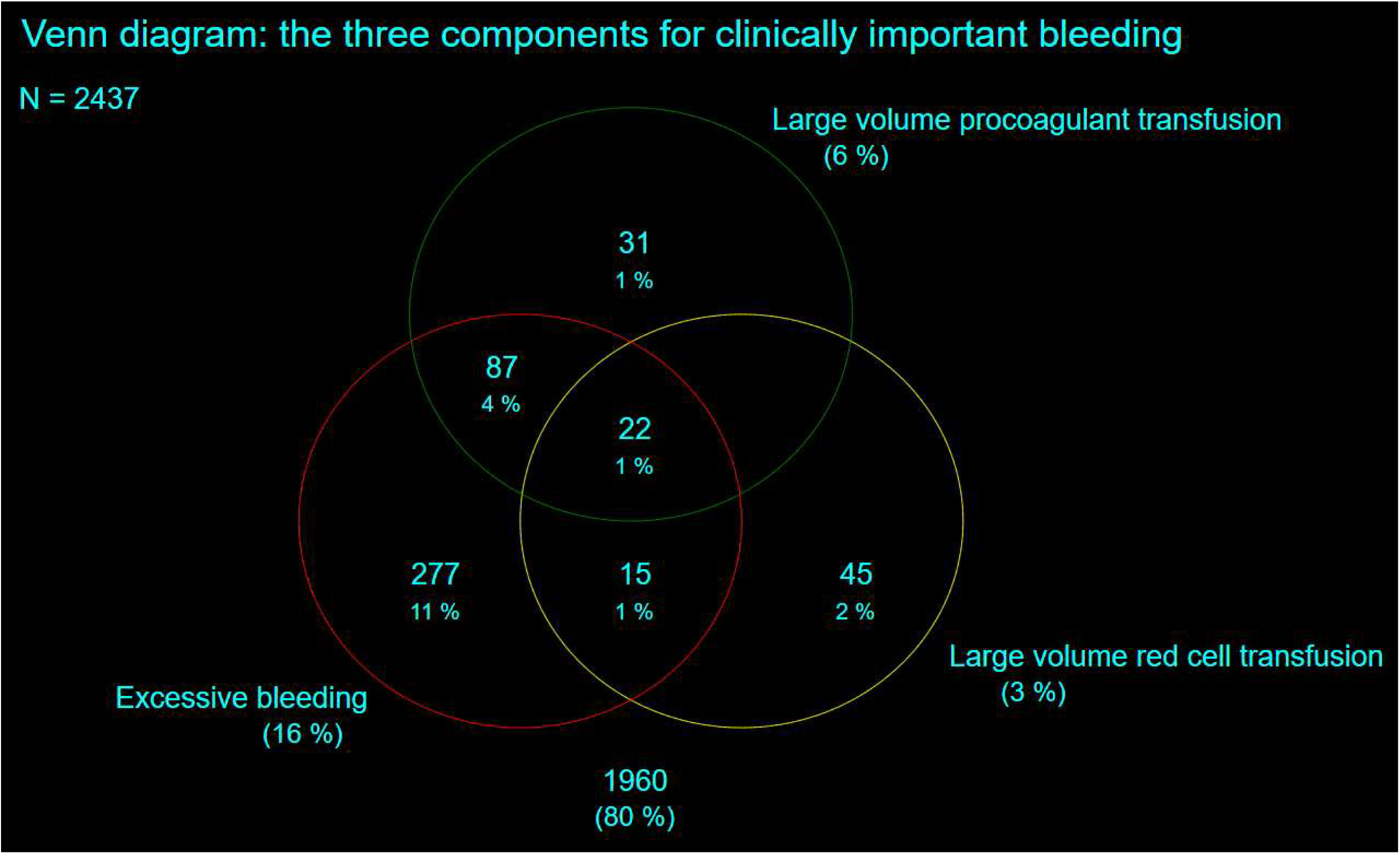
Flowchart and diagram of the study. A. Flowchart of the patient journey, the timings of diagnostic tests, and the primary and secondary clinical outcomes. B. Venn diagram of the three components for clinically important bleeding

Assessment of the primary outcome was restricted to after the routine administration of protamine before chest closure, and within 24 hours of admission to the intensive care unit. Assessment of secondary outcomes was from protamine administration until hospital discharge or death.

Decisions about intra- and post-operative haemostasis and transfusion treatment were guided by existing unit protocols and *ad hoc* TEG analyses performed without knowledge of the TEG/ROTEM/Multiplate measured remotely, and standard laboratory screening tests of coagulation (PT, APTT, ACT, fibrinogen, platelet count), performed at the discretion of the responsible clinician.

### Statistical analyses

We checked for outliers and missing values before any statistical analyses to ensure data quality. Normalisation or transformation of the biomarkers was conducted when necessary. We reported descriptive statistics by patients with and without clinically important bleeding, using mean and standard deviation (SD) or quartiles (median, Q1, Q3) as appropriate for continuous variables, and frequency (n) and percentage (%) for binary and categorical variables. Multiple imputation with chained equations was used to impute missing values for laboratory results using predictive mean matching for continuous variables with 10 imputed datasets. Results were combined using Rubin’s rules. [11] We investigated the association between individual variables and CIB using univariable logistic regression and reported the odds ratio (OR) and 95% confidence interval (95% CI), with the P value.

We investigated the diagnostic accuracy of individual parameters of TEG, ROTEM, pre-surgery organ dysfunction biomarkers, biological ageing biomarkers, post-protamine laboratory tests of coagulation, multiplate aggregometry, and Sysmex for CIB and its three components and reported area under the receiver operating characteristic curve (AUROC) and 95% CI. We compared the AUROC values between the continuous and binary forms for each parameter/biomarker, where the binary form was based on pre-specified thresholds (**eTables A to D**) to dichotomise the continuous values into normal and abnormal test results. Where cut-offs for biomarkers were not well-established, only continuous values were used in the analyses. Sensitivity analysis was conducted when restricted to participants with clinical concern about bleeding, as defined in the primary COPTIC analysis.

Different modelling strategies were used to predict CIB and its three components. When using variables from multiple test domains, two machine learning methods, stepwise backward elimination and Lasso (least absolute shrinkage and selection operator) logistic regression were used to guide variable selection in the model development stage in the imputed datasets, which was an iterative process. We retained variables with statistical significance (P<0.05) in the multi-variable models. We balanced model parsimony and clinical utility when selecting the best set of predictors for the final model. A simple model with reasonably satisfactory model performance and the potential to translate into clinical practice, is preferred to complicated models with more variables. The models were then internally validated using 10-fold cross-validation. Model performance was evaluated by AUROC and 95% CI for discrimination (whether an individual would have a positive binary outcome or not), Observed-to-Expected (O:E) Ratio and Brier score for calibration (the agreement between the observed frequencies and the predictive probabilities of the outcome), and net benefit for clinical utility (balance the true positives and false positive across probability thresholds) using decision curve analysis. [12] AUROC values were interpreted as poor <0.7, moderate 0.7-0.8, good 0.8-0.9, and excellent >0.9. [13, 14] Models with Brier scores close to 0 and O:E ratio close to 1 have better calibration. All analyses were conducted in Stata 18.

## Results

### Analysis cohort and the prevalence of primary and secondary outcomes

COPTIC participants included (n=2437) and excluded (n=38, 1.5%) from this study were similar for all baseline clinical variables except for age, which was younger in the excluded cohort (**eTable 1**). In the analysis population, participants who developed clinically important bleeding 477/2437 (19.6%) were more likely to be older, male, have lower BMI, and moderate or poor left ventricular function, anaemia, peripheral vascular disease, have surgery other than isolated CABG, and longer cardiopulmonary bypass times (**Table 1**). Clinically important bleeding increased the likelihood of developing a composite secondary outcome of MI, stroke, Stage 3 AKI, sepsis, or death; OR=2.3 (95%CI 1.85-2.92).

**Table 1.**
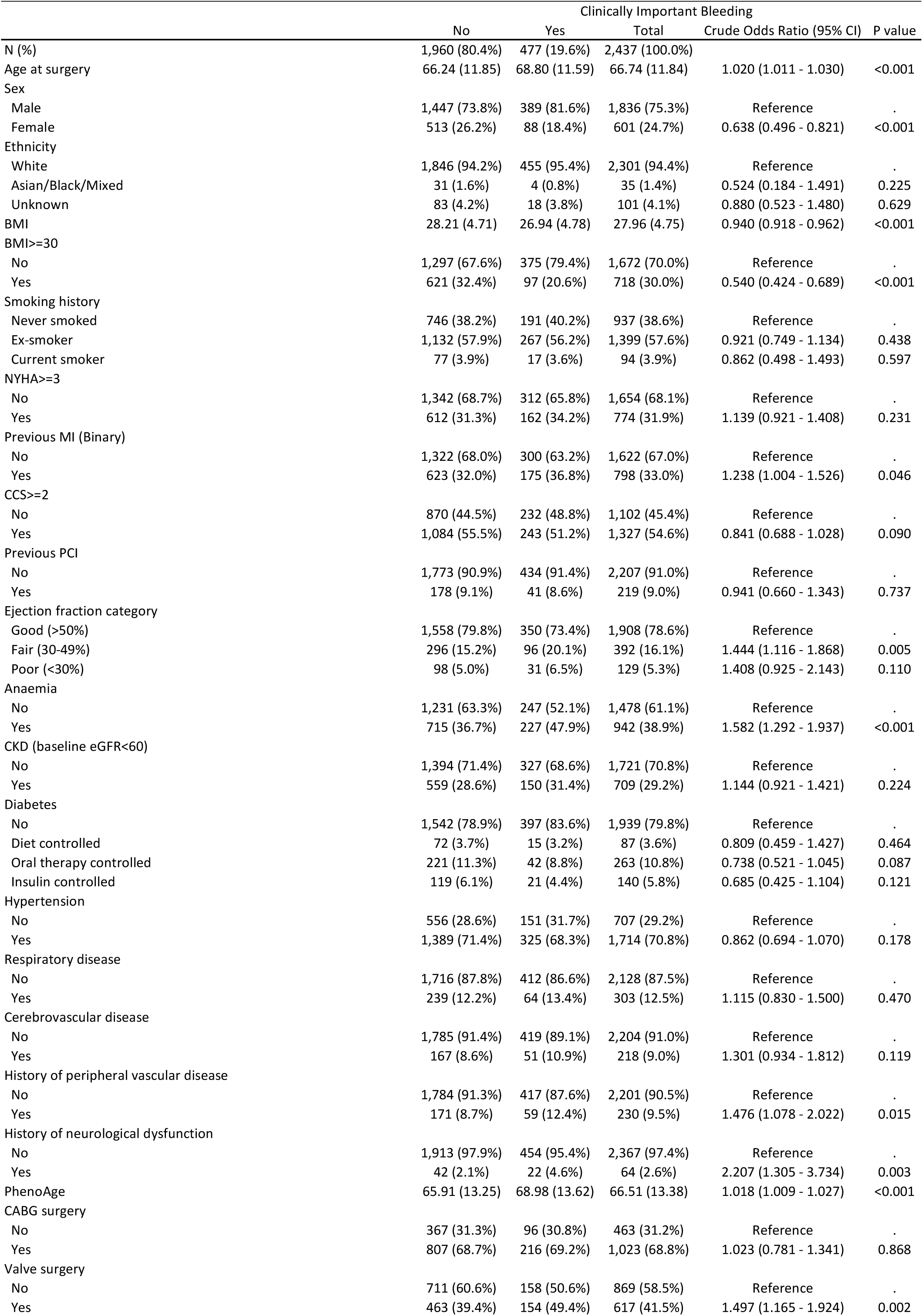
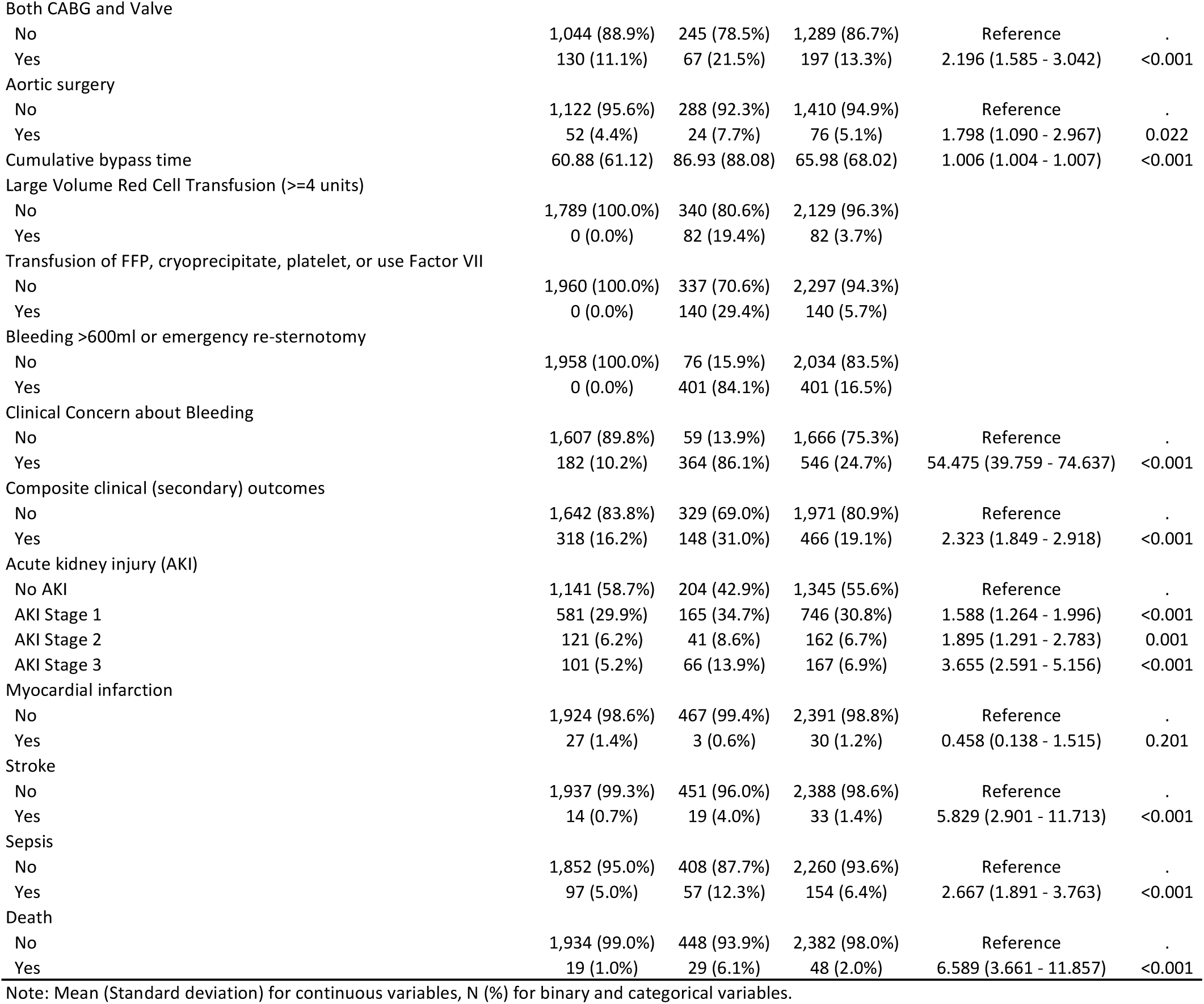
Baseline clinical characteristics in participants with and without clinically important bleeding.

|  | Clinically Important Bleeding |  |  | Crude Odds Ratio (95% CI) | P value |
| --- | --- | --- | --- | --- | --- |
|  | No | Yes | Total |  |  |
| N (%) | 1,960 (80.4%) | 477 (19.6%) | 2,437 (100.0%) |  |  |
| Age at surgery | 66.24 (11.85) | 68.80 (11.59) | 66.74 (11.84) | 1.020 (1.011 - 1.030) | <0.001 |
| Sex |  |  |  |  |  |
| Male | 1,447 (73.8%) | 389 (81.6%) | 1,836 (75.3%) | Reference | . |
| Female | 513 (26.2%) | 88 (18.4%) | 601 (24.7%) | 0.638 (0.496 - 0.821) | <0.001 |
| Ethnicity |  |  |  |  |  |
| White | 1,846 (94.2%) | 455 (95.4%) | 2,301 (94.4%) | Reference | . |
| Asian/Black/Mixed | 31 (1.6%) | 4 (0.8%) | 35 (1.4%) | 0.524 (0.184 - 1.491) | 0.225 |
| Unknown | 83 (4.2%) | 18 (3.8%) | 101 (4.1%) | 0.880 (0.523 - 1.480) | 0.629 |
| BMI | 28.21 (4.71) | 26.94 (4.78) | 27.96 (4.75) | 0.940 (0.918 - 0.962) | <0.001 |
| BMI>=30 |  |  |  |  |  |
| No | 1,297 (67.6%) | 375 (79.4%) | 1,672 (70.0%) | Reference | . |
| Yes | 621 (32.4%) | 97 (20.6%) | 718 (30.0%) | 0.540 (0.424 - 0.689) | <0.001 |
| Smoking history |  |  |  |  |  |
| Never smoked | 746 (38.2%) | 191 (40.2%) | 937 (38.6%) | Reference | . |
| Ex-smoker | 1,132 (57.9%) | 267 (56.2%) | 1,399 (57.6%) | 0.921 (0.749 - 1.134) | 0.438 |
| Current smoker | 77 (3.9%) | 17 (3.6%) | 94 (3.9%) | 0.862 (0.498 - 1.493) | 0.597 |
| NYHA>=3 |  |  |  |  |  |
| No | 1,342 (68.7%) | 312 (65.8%) | 1,654 (68.1%) | Reference | . |
| Yes | 612 (31.3%) | 162 (34.2%) | 774 (31.9%) | 1.139 (0.921 - 1.408) | 0.231 |
| Previous MI (Binary) |  |  |  |  |  |
| No | 1,322 (68.0%) | 300 (63.2%) | 1,622 (67.0%) | Reference | . |
| Yes | 623 (32.0%) | 175 (36.8%) | 798 (33.0%) | 1.238 (1.004 - 1.526) | 0.046 |
| CCS>=2 |  |  |  |  |  |
| No | 870 (44.5%) | 232 (48.8%) | 1,102 (45.4%) | Reference | . |
| Yes | 1,084 (55.5%) | 243 (51.2%) | 1,327 (54.6%) | 0.841 (0.688 - 1.028) | 0.090 |
| Previous PCI |  |  |  |  |  |
| No | 1,773 (90.9%) | 434 (91.4%) | 2,207 (91.0%) | Reference | . |
| Yes | 178 (9.1%) | 41 (8.6%) | 219 (9.0%) | 0.941 (0.660 - 1.343) | 0.737 |
| Ejection fraction category |  |  |  |  |  |
| Good (>50%) | 1,558 (79.8%) | 350 (73.4%) | 1,908 (78.6%) | Reference | . |
| Fair (30-49%) | 296 (15.2%) | 96 (20.1%) | 392 (16.1%) | 1.444 (1.116 - 1.868) | 0.005 |
| Poor (<30%) | 98 (5.0%) | 31 (6.5%) | 129 (5.3%) | 1.408 (0.925 - 2.143) | 0.110 |
| Anaemia |  |  |  |  |  |
| No | 1,231 (63.3%) | 247 (52.1%) | 1,478 (61.1%) | Reference | . |
| Yes | 715 (36.7%) | 227 (47.9%) | 942 (38.9%) | 1.582 (1.292 - 1.937) | <0.001 |
| CKD (baseline eGFR<60) |  |  |  |  |  |
| No | 1,394 (71.4%) | 327 (68.6%) | 1,721 (70.8%) | Reference | . |
| Yes | 559 (28.6%) | 150 (31.4%) | 709 (29.2%) | 1.144 (0.921 - 1.421) | 0.224 |
| Diabetes |  |  |  |  |  |
| No | 1,542 (78.9%) | 397 (83.6%) | 1,939 (79.8%) | Reference | . |
| Diet controlled | 72 (3.7%) | 15 (3.2%) | 87 (3.6%) | 0.809 (0.459 - 1.427) | 0.464 |
| Oral therapy controlled | 221 (11.3%) | 42 (8.8%) | 263 (10.8%) | 0.738 (0.521 - 1.045) | 0.087 |
| Insulin controlled | 119 (6.1%) | 21 (4.4%) | 140 (5.8%) | 0.685 (0.425 - 1.104) | 0.121 |
| Hypertension |  |  |  |  |  |
| No | 556 (28.6%) | 151 (31.7%) | 707 (29.2%) | Reference | . |
| Yes | 1,389 (71.4%) | 325 (68.3%) | 1,714 (70.8%) | 0.862 (0.694 - 1.070) | 0.178 |
| Respiratory disease |  |  |  |  |  |
| No | 1,716 (87.8%) | 412 (86.6%) | 2,128 (87.5%) | Reference | . |
| Yes | 239 (12.2%) | 64 (13.4%) | 303 (12.5%) | 1.115 (0.830 - 1.500) | 0.470 |
| Cerebrovascular disease |  |  |  |  |  |
| No | 1,785 (91.4%) | 419 (89.1%) | 2,204 (91.0%) | Reference | . |
| Yes | 167 (8.6%) | 51 (10.9%) | 218 (9.0%) | 1.301 (0.934 - 1.812) | 0.119 |
| History of peripheral vascular disease |  |  |  |  |  |
| No | 1,784 (91.3%) | 417 (87.6%) | 2,201 (90.5%) | Reference | . |
| Yes | 171 (8.7%) | 59 (12.4%) | 230 (9.5%) | 1.476 (1.078 - 2.022) | 0.015 |
| History of neurological dysfunction |  |  |  |  |  |
| No | 1,913 (97.9%) | 454 (95.4%) | 2,367 (97.4%) | Reference | . |
| Yes | 42 (2.1%) | 22 (4.6%) | 64 (2.6%) | 2.207 (1.305 - 3.734) | 0.003 |
| PhenoAge | 65.91 (13.25) | 68.98 (13.62) | 66.51 (13.38) | 1.018 (1.009 - 1.027) | <0.001 |
| CABG surgery |  |  |  |  |  |
| No | 367 (31.3%) | 96 (30.8%) | 463 (31.2%) | Reference | . |
| Yes | 807 (68.7%) | 216 (69.2%) | 1,023 (68.8%) | 1.023 (0.781 - 1.341) | 0.868 |
| Valve surgery |  |  |  |  |  |
| No | 711 (60.6%) | 158 (50.6%) | 869 (58.5%) | Reference | . |
| Yes | 463 (39.4%) | 154 (49.4%) | 617 (41.5%) | 1.497 (1.165 - 1.924) | 0.002 |
| Both CABG and Valve |  |  |  |  |  |
| No | 1,044 (88.9%) | 245 (78.5%) | 1,289 (86.7%) | Reference | . |
| Yes | 130 (11.1%) | 67 (21.5%) | 197 (13.3%) | 2.196 (1.585 - 3.042) | <0.001 |
| Aortic surgery |  |  |  |  |  |
| No | 1,122 (95.6%) | 288 (92.3%) | 1,410 (94.9%) | Reference | . |
| Yes | 52 (4.4%) | 24 (7.7%) | 76 (5.1%) | 1.798 (1.090 - 2.967) | 0.022 |
| Cumulative bypass time | 60.88 (61.12) | 86.93 (88.08) | 65.98 (68.02) | 1.006 (1.004 - 1.007) | <0.001 |
| Large Volume Red Cell Transfusion (>=4 units) |  |  |  |  |  |
| No | 1,789 (100.0%) | 340 (80.6%) | 2,129 (96.3%) |  |  |
| Yes | 0 (0.0%) | 82 (19.4%) | 82 (3.7%) |  |  |
| Transfusion of FFP, cryoprecipitate, platelet, or use Factor VII |  |  |  |  |  |
| No | 1,960 (100.0%) | 337 (70.6%) | 2,297 (94.3%) |  |  |
| Yes | 0 (0.0%) | 140 (29.4%) | 140 (5.7%) |  |  |
| Bleeding >600ml or emergency re-sternotomy |  |  |  |  |  |
| No | 1,958 (100.0%) | 76 (15.9%) | 2,034 (83.5%) |  |  |
| Yes | 0 (0.0%) | 401 (84.1%) | 401 (16.5%) |  |  |
| Clinical Concern about Bleeding |  |  |  |  |  |
| No | 1,607 (89.8%) | 59 (13.9%) | 1,666 (75.3%) | Reference | . |
| Yes | 182 (10.2%) | 364 (86.1%) | 546 (24.7%) | 54.475 (39.759 - 74.637) | <0.001 |
| Composite clinical (secondary) outcomes |  |  |  |  |  |
| No | 1,642 (83.8%) | 329 (69.0%) | 1,971 (80.9%) | Reference | . |
| Yes | 318 (16.2%) | 148 (31.0%) | 466 (19.1%) | 2.323 (1.849 - 2.918) | <0.001 |
| Acute kidney injury (AKI) |  |  |  |  |  |
| No AKI | 1,141 (58.7%) | 204 (42.9%) | 1,345 (55.6%) | Reference | . |
| AKI Stage 1 | 581 (29.9%) | 165 (34.7%) | 746 (30.8%) | 1.588 (1.264 - 1.996) | <0.001 |
| AKI Stage 2 | 121 (6.2%) | 41 (8.6%) | 162 (6.7%) | 1.895 (1.291 - 2.783) | 0.001 |
| AKI Stage 3 | 101 (5.2%) | 66 (13.9%) | 167 (6.9%) | 3.655 (2.591 - 5.156) | <0.001 |
| Myocardial infarction |  |  |  |  |  |
| No | 1,924 (98.6%) | 467 (99.4%) | 2,391 (98.8%) | Reference | . |
| Yes | 27 (1.4%) | 3 (0.6%) | 30 (1.2%) | 0.458 (0.138 - 1.515) | 0.201 |
| Stroke |  |  |  |  |  |
| No | 1,937 (99.3%) | 451 (96.0%) | 2,388 (98.6%) | Reference | . |
| Yes | 14 (0.7%) | 19 (4.0%) | 33 (1.4%) | 5.829 (2.901 - 11.713) | <0.001 |
| Sepsis |  |  |  |  |  |
| No | 1,852 (95.0%) | 408 (87.7%) | 2,260 (93.6%) | Reference | . |
| Yes | 97 (5.0%) | 57 (12.3%) | 154 (6.4%) | 2.667 (1.891 - 3.763) | <0.001 |
| Death |  |  |  |  |  |
| No | 1,934 (99.0%) | 448 (93.9%) | 2,382 (98.0%) | Reference | . |
| Yes | 19 (1.0%) | 29 (6.1%) | 48 (2.0%) | 6.589 (3.661 - 11.857) | <0.001 |

Note: Mean (Standard deviation) for continuous variables, N (%) for binary and categorical variables.

For the individual components of the primary outcome, large volume red cell transfusion occurred in 82/2437 (5.7%) participants, large volume/dose of procoagulants occurred in 140/2437 (5.7%), and excessive bleeding occurred in 401 (16.5%). 30.9% (124/401) patients had at least two components (Venn diagram, **Figure 1B**). Test results for all biomarkers in participants with and without CIB are shown in **eTable 2**. This univariable analysis found 35 out of 60 test parameters/biomarkers (continuous) were significantly associated with CIB (P<0.05).

### Diagnostic accuracy of TEG/ROTEM for laboratory diagnostic tests and aggregometry

We assessed whether TEG/ROTEM results demonstrated diagnostic accuracy for test positive thresholds of corresponding laboratory tests of coagulopathy, full blood count, and platelet function. When the threshold value for a positive test for each TEG parameter was used as binary variables in the analysis, all the AUROC values were <0.6 (**eTable 3**). When continuous TEG parameters were used, the majority of AUROC values were <0.7, except for TEG Citrate Kaolin (CK) R Time (rmin) for Haemoglobin 0.718 (0.641–0.795) and CK Heparinase rmin for Anti-Xa activity 0.852 (0.672–1.000). TEG CK Maximum Amplitude (MA) demonstrated moderate (AUROC<0.8) predictive accuracy for Factor XIII, vWF, and post-op platelet counts, and good predictive accuracy for Clauss Fibrinogen 0.829 (0.812–0.847), and the product of platelet count and mean platelet volume (MPV X platelet), 0.808 (0.789–0.827).

When the threshold value for a positive test for each ROTEM parameter was used (binary variables), all the resulting AUROC values were <0.7, except for INTEM α angle and MPV X platelet, 0.703 (0.684–0.721, **eTable 4**). When continuous ROTEM parameters were used, the AUROC for EXTEM Clotting Time (CT) for PT was 0.788 (0.731–0.845), HEPTEM CT for Anti-Xa was 0.765 (0.609–0.921), INTEM α angle for Clauss Fibrinogen was 0.848 (0.831–0.864), and 0.848 (0.832–0.864) for platelet count. FIBTEM Maximum Clot Firmness (MCF) demonstrated AUROC 0.908 (0.895–0.920) for Clauss Fibrinogen, and INTEM MCF demonstrated AUROC 0.8-0.9 for Claus Fibrinogen, Platelet count, and MPV X Platelet.

### Predictive accuracy of TEG/ ROTEM and laboratory tests for primary and secondary outcomes

The results of AUROC for individual TEG/ROTEM parameters for the primary outcome (CIB), three individual components of the primary outcome, and secondary outcomes in univariate analysis are in **Table 2**. All AUROC values were <0.7, regardless of input of TEG/ROTEM parameters as continuous values or using the established positive test thresholds as binary variables.

**Table 2.**
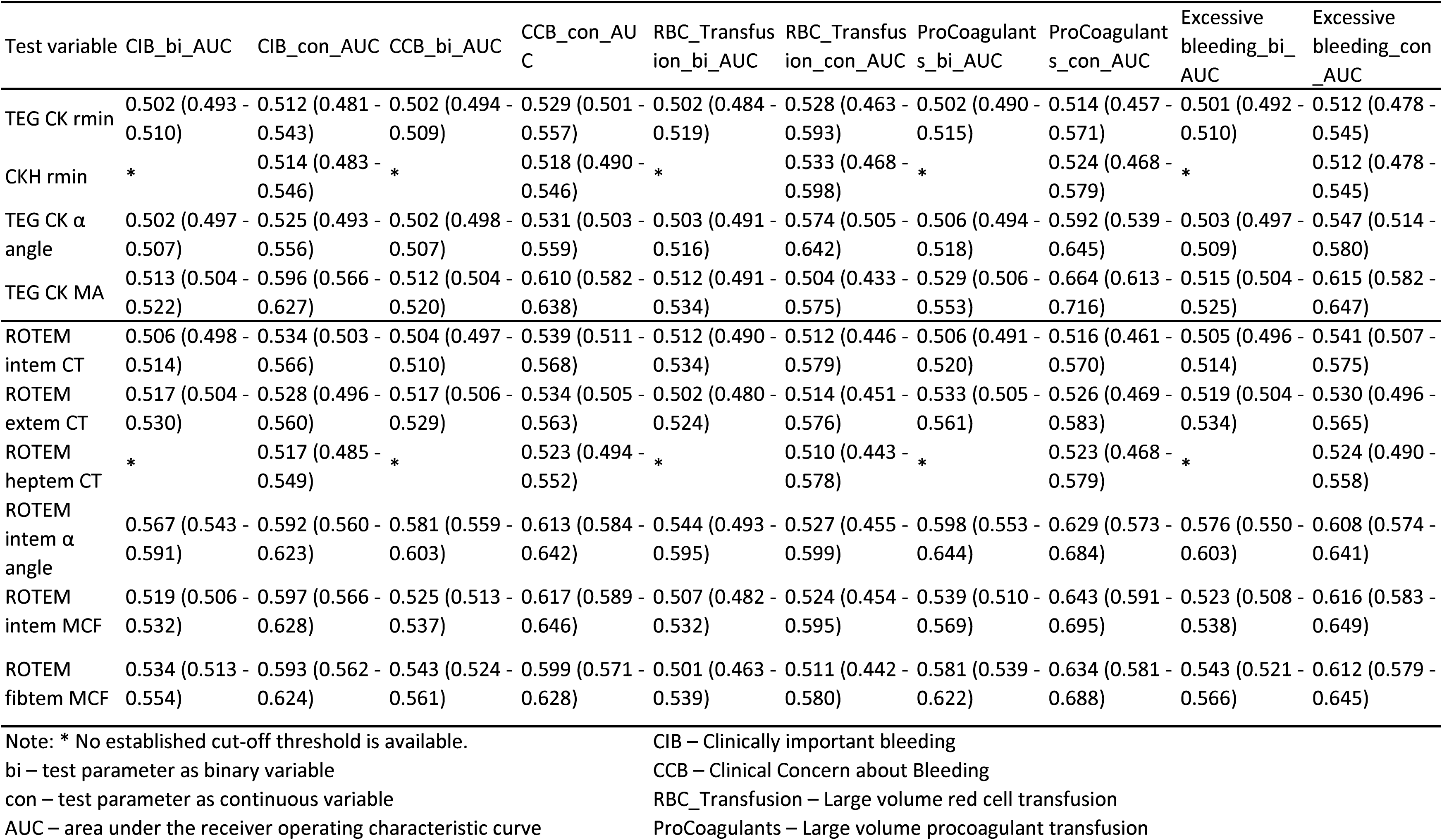
Predictive accuracy of TEG/ROTEM parameters (continuous and binary) for clinically important bleeding and its three individual components, and the secondary outcome.

**Table 3.** Area under the receiver operating characteristic curve (AUROC, discrimination of prediction models) for clinically important bleeding and its individual components.

| Models | Clinically important bleeding | Large volume red cell transfusion | Large volume procoagulant transfusion | Excessive bleeding |
| --- | --- | --- | --- | --- |
| TEG | 0.611 (0.521 - 0.701) | * | 0.649 (0.501 - 0.798) | 0.620 (0.523 - 0.716) |
| ROTEM | 0.588 (0.496 - 0.681) | 0.580 (0.392 - 0.766) | 0.630 (0.487 - 0.774) | 0.606 (0.508 - 0.704) |
| Organ dysfunction biomarkers | 0.606 (0.512 - 0.701) | **0.701 (0.501 - 0.900)** | 0.651 (0.497 - 0.805) | 0.602 (0.501 - 0.702) |
| Ageing biomarkers | 0.536 (0.433 - 0.639) | 0.646 (0.410 - 0.880) | * | 0.530 (0.419 - 0.641) |
| Lab tests: post-op (Sysmex + Multiplate + Coagulation) | 0.688 (0.607 - 0.769) | **0.745 (0.602 - 0.886)** | **0.720 (0.595 - 0.845)** | 0.680 (0.592 - 0.769) |
| Organ dysfunction biomarkers + Ageing biomarkers + Lab tests | **0.701 (0.620 - 0.781)** | **0.754 (0.602 - 0.903)** | **0.723 (0.590 - 0.857)** | **0.701 (0.613 - 0.788)** |
| TEG + Organ dysfunction biomarkers + Ageing biomarkers + Lab tests | 0.694 (0.612 - 0.775) | 0.762 (0.626 - 0.897) | 0.732 (0.606 - 0.858) | 0.686 (0.596 - 0.776) |
| ROTEM + Organ dysfunction biomarkers + Ageing biomarkers + Lab tests | 0.692 (0.610 - 0.775) | **0.753 (0.608 - 0.895)** | **0.732 (0.606 - 0.858)** | 0.695 (0.606 - 0.783) |

Note: \* No significant predictors for the model

The results of AUROC for individual laboratory reference tests of coagulation, full blood count, aggregometry, organ dysfunction, and biological ageing biomarkers for the outcomes in univariate analyses are shown in **eTable 5**. All AUROC values were <0.7, regardless of input as continuous values or as binary variables based on the published positive test thresholds, except for post-protamine haemoglobin 0.723 (0.670–0.776) and haematocrit 0.723 (0.670–0.776) for large volume red cell transfusion. Sensitivity analyses restricted to participants who developed clinical concern about bleeding to actively bleeding patients, did not alter the overall conclusion.

### Best predictive models using all test parameters

Multivariable models with different combinations of biomarkers and test parameters to predict the primary outcome and its three components were developed and internally validated using 10-fold cross-validation. Predictors in each model for the outcomes are listed in **eTable 6**. ROC curves showing model discrimination and decision curve analysis showing the clinical utility of different models for the primary outcome (A) and its three individual components (B-D) are shown in **Figures 2 and 3**, respectively. The Brier scores were close among the models for each outcome, and the O:E ratios were close to 1, which indicated good calibration (both in **eTable 6**), meaning the predicted probabilities had high agreement with the observed frequencies.

**Figure 2.**
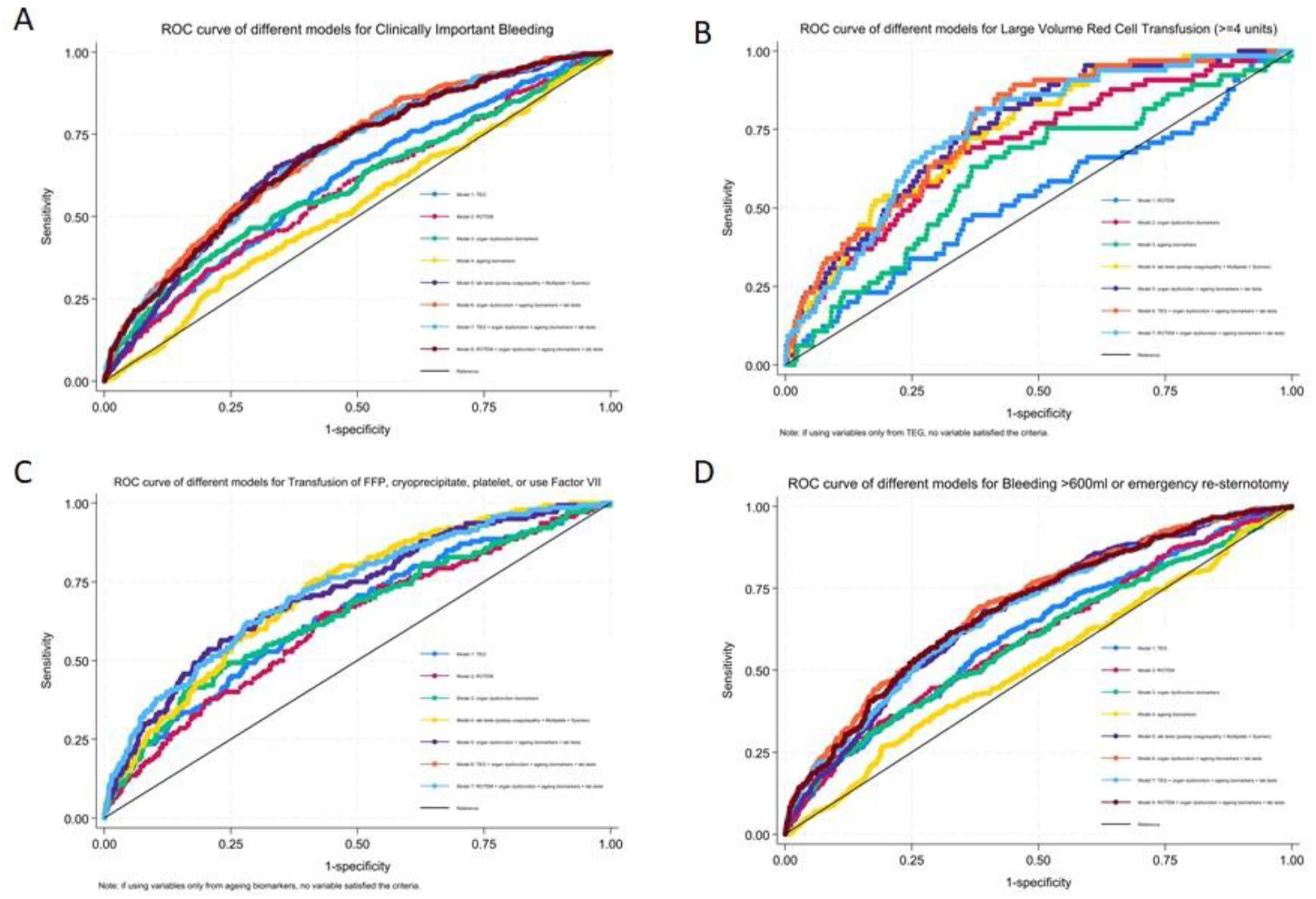
ROC curve showing model discrimination for clinically important bleeding and its three individual components.

**Figure 3.**
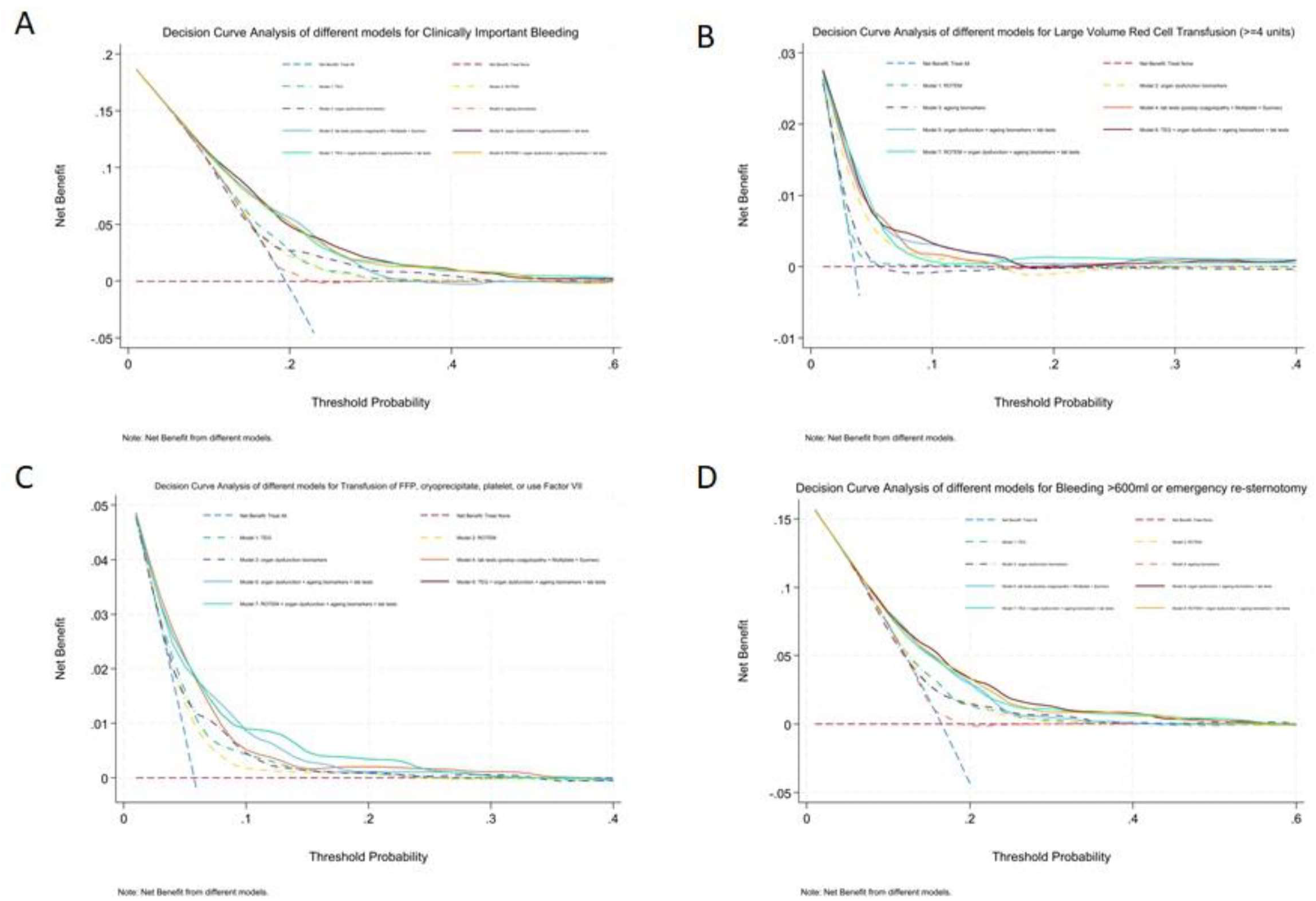
Decision curve analysis for clinically important bleeding and its three individual components.

Models using the predictors from TEG or ROTEM had lower AUROC values (<0.7) for the primary outcome and its individual components, and worse clinical utility. The model derived from full blood count, aggregometry and laboratory tests of coagulation demonstrated AUROC 0.688 (0.607–0.769) for the primary outcome, AUROC 0.745 (0.602–0.886) for large volume red cell transfusion, AUROC 0.720 (0.595–0.845) for large volume procoagulant transfusion, and AUROC 0.680 (0.592–0.769) for excessive bleeding.

With additional organ dysfunction biomarkers and ageing biomarkers to the laboratory tests, the AUROC values improved slightly for the primary outcome 0.701 (0.620–0.781), large volume red cell transfusion 0.754 (0.602–0.903), large volume procoagulant transfusion 0.723 (0.590–0.857), and excessive bleeding 0.701 (0.613–0.788), but had similar clinical utility. TEG or ROTEM parameters were not selected as predictors in the final multivariable models.

## Discussion

### Main findings

TEG demonstrated moderate to good predictive accuracy, and ROTEM shown good to excellent predictive accuracy for laboratory reference tests of coagulation, low haematocrit, and low platelet counts/volume product, but were poor predictors for clinically important bleeding. FBC, Multiplate, and laboratory tests of coagulation also demonstrated poor predictive accuracy for clinically important bleeding. The addition of multimorbidity and biological ageing biomarker results did not significantly improve predictive accuracy for clinically important bleeding beyond that observed with the laboratory data.

TEG/ROTEM demonstrated poor predictive accuracy for all three components of the primary outcome. Multiplate, coagulation tests, and FBC data demonstrated moderate predictive accuracy for both large volume red cell transfusion and large volume procoagulant transfusion, but not for excessive bleeding. Additional multimorbidity and biological ageing biomarkers yielded small increases in predictive accuracy with similar clinical utility, with the largest AUROC of 0.701 for excessive bleeding among all the prediction models.

### Strengths and limitations

COPTIC remains the largest diagnostic test accuracy study of bleeding tests in cardiac surgery. [7] The cohort is unique as it provides simultaneous measurement of multiple disease processes associated with bleeding using high quality prospectively collected clinical data and standardised assays performed in a dedicated laboratory. We used the QUADAS framework to minimise potential bias. Specifically, the study design avoided the limitations of previous similar studies identified in a systematic review, including non-prespecified test thresholds, non-blinded clinical staff, small sample sizes, and limited applicability. [7] We used regularisation techniques (Lasso) to inform the selection of predictors from several dozens of biomarkers/test parameters, to get simpler models with fewer parameters for better interpretability and for potential clinical use. We also avoided using “black box” algorithms or yielding overly complex models that may just capture the noise of the data. In addition, we used 10-fold cross-validation to internally validate the models, and conducted a comprehensive assessment of model performance for discrimination, calibration, and clinical utility.

The first limitation is the age of the study. COPTIC recruited participants between 2010 and 2012, and cardiac surgery populations have changed over the last decade. However, this is mitigated by the sample size, including over 90% of all elective and urgent patients operated on in a single large tertiary cardiac centre over 2 years. This included many high-risk procedures, typical of contemporary practice, and minimised applicability bias. We consider degradation of stored plasma samples unlikely; baseline biomarker values for multimorbidity and ageing in the COPTIC samples were comparable to those from more recent cohorts. [8] Second, the study evaluates the TEG® 5000 Thromboelastograph® that has been superseded by the TEG® 6s Hemostasis Analyser. In mitigation, the principles underlying both platforms are similar. Essentially, the mechanical attributes of a clot are used to infer specific defects in blood clotting, and the observed differences in predictive accuracy between the two platforms are not clinically significant [15, 16]. The TEG findings were also replicated with the ROTEM device which demonstrated better predictive accuracy for individual defects in clotting, platelet aggregation and haematopoiesis than the TEG 5000®.

### Clinical interpretation

First, the study shows that the measurement of blood derived factors involved in clotting could not predict clinically important bleeding well, specifically excessive bleeding, despite using a data-driven machine learning approach to select the most relevant predictors. One potential explanation, based on Virchow’s triad, is that the contribution of the vessel wall may represent an important unmeasured determinant of bleeding. Preliminary evidence to support this comes from a recent study where we showed that a patient cluster with increased bleeding had significant reductions in ICAM-1 levels in plasma, indicative of altered vascular activation [8]. Vascular dysfunction also underlies organ injury following cardiopulmonary bypass [17], providing a new hypothesis for the bleeding paradox for future research.

Second, these observations lead us to hypothesise that the three components of the primary outcome represent different mechanisms. This is supported by the Venn diagram, and the differences in the biomarkers showing various predictive accuracy for each component. This suggests that composite definitions of clinically important bleeding in this and previous studies [5, 6], or the use of the individual components interchangeably [1, 2], are likely barriers to understanding the underlying processes. This information will assist in further attempts to identify mechanisms or phenotypes underlying bleeding, and organ dysfunction, and enable the development of targeted treatments.

### Conclusion

Current diagnostic tests demonstrate moderate predictive accuracy for excessive bleeding following cardiac surgery. Additional biomarkers of multimorbidity, organ dysfunction, and biological ageing did not improve discrimination or clinical utility substantially. Revised definitions of excessive bleeding phenotypes based on these results should reduce barriers to the understanding of underlying disease processes.

### Funders

GJM, WL, HA, YL, and MW are funded by British Heart Foundation CH/12/1/29419. The COPTIC Study was funded by the National Institute for Health and Care Research (NIHR, ref: RP-PG-0407-10384). RG is funded by the Leicester NIHR Biomedical Research Centre (ref: NIHR203327), and the biomarker assays were part funded by this grant, along with the British Heart Foundation Leicester Accelerator Award AA/18/3/34220, and British Heart Foundation Grant RG/17/9/32812. LG is funded by NHS Blood and Transplant. The funders have no role in data collection, analysis, interpretation of findings, writing the manuscript, or the decision to submit and publish this article.

## Supporting information

COPTIC-C. Supplementary materials

## Data Availability

For potential collaborators, please contact the chief investigator to discuss.

## Acknowledgements

We would like to thank the participants contributing to the COPTIC study, and the study team at University Hospitals Bristol and Weston NHS Foundation Trust.

## Contributions from study authors

GJM and WL designed the study. WL performed the statistical analyses. GJM and WL interpreted the results and drafted the manuscript. GJM is the senior author and guarantor of the study. AM designed the COPTIC study, and was the chief investigator of the COPTIC A, B and COPTIC LRT studies and led the laboratory analyses of the data subsequently used in this study. RG and LG co-designed the study and interpreted the study data. MW designed and supervised the laboratory analyses of multimorbidity and biological ageing biomarkers. HP undertook the laboratory analyses. FL performed some of the statistical analyses. HA was the coordinator of the study. All authors approved the final manuscript for submission.

## eTables for Results (published as supplementary materials)

**eTable 1.** Characteristics of the COPTIC participants who were included or excluded from the analysis dataset.

|  | Included in analysis | Patients in the COPTIC study |  |
| --- | --- | --- | --- |
|  |  | Consented patients excluded from analysis | P value |
| N (%) | 2,437 (98.5%) | 38 (1.5%) |  |
| Age at surgery | 66.74 (11.84) | 61.19 (14.67) | 0.005 |
| Sex |  |  |  |
| Male | 1,836 (75.3%) | 29 (76.3%) | 0.890 |
| Female | 601 (24.7%) | 9 (23.7%) |  |
| Ethnicity |  |  |  |
| White | 2,301 (94.4%) | 35 (97.2%) | 0.702 |
| Asian/Black/Mixed | 35 (1.4%) | 0 (0.0%) |  |
| Unknown | 101 (4.1%) | 1 (2.8%) |  |
| BMI | 27.96 (4.75) | 27.34 (5.55) | 0.446 |
| BMI>=30 |  |  |  |
| No | 1,672 (70.0%) | 26 (74.3%) | 0.579 |
| Yes | 718 (30.0%) | 9 (25.7%) |  |
| Smoking history |  |  |  |
| Never smoked | 937 (38.6%) | 11 (31.4%) | 0.630 |
| Ex-smoker | 1,399 (57.6%) | 22 (62.9%) |  |
| Current smoker | 94 (3.9%) | 2 (5.7%) |  |
| NYHA>=3 |  |  |  |
| No | 1,654 (68.1%) | 27 (77.1%) | 0.255 |
| Yes | 774 (31.9%) | 8 (22.9%) |  |
| Previous MI (Binary) |  |  |  |
| No | 1,622 (67.0%) | 22 (62.9%) | 0.603 |
| Yes | 798 (33.0%) | 13 (37.1%) |  |
| CCS>=2 |  |  |  |
| No | 1,102 (45.4%) | 16 (45.7%) | 0.967 |
| Yes | 1,327 (54.6%) | 19 (54.3%) |  |
| Previous PCI |  |  |  |
| No | 2,207 (91.0%) | 33 (94.3%) | 0.496 |
| Yes | 219 (9.0%) | 2 (5.7%) |  |
| Ejection fraction category |  |  |  |
| Good (>50%) | 1,908 (78.6%) | 28 (77.8%) | 0.698 |
| Fair (30-49%) | 392 (16.1%) | 5 (13.9%) |  |
| Poor (<30%) | 129 (5.3%) | 3 (8.3%) |  |
| Anaemia |  |  |  |
| No | 1,478 (61.1%) | 22 (62.9%) | 0.830 |
| Yes | 942 (38.9%) | 13 (37.1%) |  |
| CKD (baseline eGFR<60) |  |  |  |
| No | 1,721 (70.8%) | 0 (0.0%) | 0.119 |
| Yes | 709 (29.2%) | 1 (100.0%) |  |
| Diabetes |  |  |  |
| No | 1,939 (79.8%) | 32 (91.4%) | 0.275 |
| Diet controlled | 87 (3.6%) | 0 (0.0%) |  |
| Oral therapy controlled | 263 (10.8%) | 1 (2.9%) |  |
| Insulin controlled | 140 (5.8%) | 2 (5.7%) |  |
| Hypertension |  |  |  |
| No | 707 (29.2%) | 13 (37.1%) | 0.306 |
| Yes | 1,714 (70.8%) | 22 (62.9%) |  |
| Respiratory disease |  |  |  |
| No | 2,128 (87.5%) | 30 (85.7%) | 0.746 |
| Yes | 303 (12.5%) | 5 (14.3%) |  |
| Cerebrovascular disease |  |  |  |
| No | 2,204 (91.0%) | 33 (94.3%) | 0.499 |
| Yes | 218 (9.0%) | 2 (5.7%) |  |
| History of peripheral vascular disease |  |  |  |
| No | 2,201 (90.5%) | 32 (91.4%) | 0.858 |
| Yes | 230 (9.5%) | 3 (8.6%) |  |
| History of neurological dysfunction |  |  |  |
| No | 2,367 (97.4%) | 33 (94.3%) | 0.262 |
| Yes | 64 (2.6%) | 2 (5.7%) |  |
| Acute kidney injury (AKI) |  |  |  |
| No AKI | 1,345 (55.6%) | 0 (0.0%) | 0.004 |
| AKI Stage 1 | 746 (30.8%) | 0 (0.0%) |  |
| AKI Stage 2 | 162 (6.7%) | 0 (0.0%) |  |
| AKI Stage 3 | 167 (6.9%) | 1 (100.0%) |  |
| PhenoAge | 66.51 (13.38) | . (.) | . |
| CABG surgery |  |  |  |
| No | 463 (31.2%) | 8 (38.1%) | 0.496 |
| Yes | 1,023 (68.8%) | 13 (61.9%) |  |
| Valve surgery |  |  |  |
| No | 869 (58.5%) | 14 (66.7%) | 0.449 |
| Yes | 617 (41.5%) | 7 (33.3%) |  |
| Both CABG and Valve |  |  |  |
| No | 1,289 (86.7%) | 20 (95.2%) | 0.252 |
| Yes | 197 (13.3%) | 1 (4.8%) |  |
| Aortic surgery |  |  |  |
| No | 1,410 (94.9%) | 20 (95.2%) | 0.942 |
| Yes | 76 (5.1%) | 1 (4.8%) |  |
| Cumulative bypass time | 65.98 (68.02) | 58.20 (60.13) | 0.501 |

Note: Mean (Standard deviation) for continuous variables, N (%) for binary and categorical variables.

**eTable 2.** Descriptive statistics and uni-variable logistic regression of tests in participants with and without clinically important bleeding. Note: tests include TEG, ROTEM, laboratory tests of coagulation, Sysmex full blood count, multiplate aggregometry, baseline biomarkers of multimorbidity and biological ageing

|  | Clinically Important Bleeding |  |  | Odds Ratio (95% CI) | P value |
| --- | --- | --- | --- | --- | --- |
|  | No | Yes | Total |  |  |
| N (%) | 1,960 (80.4%) | 477 (19.6%) | 2,437 (100.0%) |  |  |
| TEG CK rmin (con) | 6.18 (1.95) | 6.21 (2.31) | 6.18 (2.02) | 1.01 (0.96 - 1.06) | 0.782 |
| CKH rmin (con) | 5.90 (1.37) | 5.85 (1.38) | 5.89 (1.37) | 0.97 (0.90 - 1.05) | 0.521 |
| TEG CK α angle (con) | 62.59 (7.18) | 61.84 (7.76) | 62.45 (7.30) | 0.99 (0.97 - 1.00) | 0.062 |
| TEG CK MA (con) | 59.18 (6.43) | 56.84 (6.92) | 58.74 (6.59) | 0.95 (0.93 - 0.96) | <0.001 |
| ROTEM intem CT (con) | 169.14 (34.85) | 173.14 (32.07) | 169.90 (34.37) | 1.00 (1.00 - 1.01) | 0.048 |
| ROTEM extem CT (con) | 57.49 (11.82) | 59.23 (13.75) | 57.82 (12.22) | 1.01 (1.00 - 1.02) | 0.010 |
| ROTEM heptem CT (con) | 168.63 (28.60) | 171.13 (31.75) | 169.10 (29.23) | 1.00 (1.00 - 1.01) | 0.123 |
| ROTEM intem α angle (con) | 73.77 (5.00) | 71.76 (6.34) | 73.39 (5.34) | 0.94 (0.92 - 0.96) | <0.001 |
| ROTEM intem MCF (con) | 61.60 (6.15) | 59.31 (6.77) | 61.17 (6.34) | 0.94 (0.93 - 0.96) | <0.001 |
| ROTEM fibtem MCF (con) | 14.22 (5.38) | 12.77 (5.47) | 13.95 (5.42) | 0.95 (0.92 - 0.97) | <0.001 |
| post-op HB (con) | 10.06 (1.64) | 9.33 (1.46) | 9.92 (1.63) | 0.74 (0.69 - 0.80) | <0.001 |
| post-op Haematocrit (con) | 0.29 (0.05) | 0.27 (0.04) | 0.28 (0.05) | 0.00 (0.00 - 0.00) | <0.001 |
| post-op lymphocyte count (con) | 1.47 (1.85) | 1.25 (1.50) | 1.42 (1.79) | 0.74 (0.63 - 0.88) | 0.001 |
| post-op monocyte count (con) | 0.34 (0.28) | 0.34 (0.94) | 0.34 (0.48) | 0.96 (0.73 - 1.25) | 0.753 |
| post-op neutrophil/lymphocyte Ratio (con) | 6.48 (4.60) | 7.62 (5.58) | 6.70 (4.82) | 1.04 (1.02 - 1.07) | <0.001 |
| post-op mean corpuscular volume (con) | 85.59 (5.01) | 85.95 (5.22) | 85.66 (5.05) | 1.01 (0.99 - 1.04) | 0.183 |
| post-op mean corpuscular haemoglobin (con) | 30.05 (1.82) | 30.28 (1.89) | 30.09 (1.83) | 1.07 (1.01 - 1.14) | 0.020 |
| post-op Absolute reticulocytes (con) | 41.84 (17.02) | 39.19 (17.50) | 41.33 (17.14) | 0.99 (0.98 - 1.00) | 0.004 |
| post-op Platelets (con) | 149.21 (51.22) | 133.01 (45.77) | 146.13 (50.62) | 0.99 (0.99 - 1.00) | <0.001 |
| post-op MPV (con) | 10.44 (0.99) | 10.29 (0.97) | 10.41 (0.99) | 0.85 (0.76 - 0.95) | 0.004 |
| post-op IPF (con) | 3.58 (2.29) | 3.56 (2.26) | 3.58 (2.28) | 0.99 (0.95 - 1.04) | 0.824 |
| Post-op MPV X Platelets (con) | 1,547.51 (511.75) | 1,361.36 (457.91) | 1,512.04 (507.14) | 1.00 (1.00 - 1.00) | <0.001 |
| Post-op PT (con) | 12.43 (1.11) | 12.79 (1.32) | 12.50 (1.16) | 1.28 (1.17 - 1.39) | <0.001 |
| Post-op APTT (con) | 32.22 (6.80) | 34.75 (9.88) | 32.72 (7.58) | 1.04 (1.02 - 1.06) | <0.001 |
| Anti-Xa heparin (con) | 0.19 (0.14) | 0.20 (0.16) | 0.20 (0.14) | 1.31 (0.64 - 2.68) | 0.467 |
| Endogenous thrombin potential (con) | 1,044.43 (479.82) | 991.55 (460.06) | 1,033.87 (476.30) | 1.00 (1.00 - 1.00) | 0.042 |
| Clauss Fibrinogen (con) | 2.24 (0.77) | 1.93 (0.72) | 2.18 (0.77) | 0.53 (0.45 - 0.63) | <0.001 |
| D dimer (con) | 691.16 (1,023.28) | 881.20 (1,277.26) | 728.91 (1,080.84) | 1.00 (1.00 - 1.00) | 0.002 |
| Factor XIII (con) | 81.58 (20.30) | 74.78 (19.08) | 80.23 (20.24) | 0.98 (0.98 - 0.99) | <0.001 |
| Von Willebrand factor ristocetin cofactor (con) | 159.02 (59.32) | 161.31 (68.94) | 159.48 (61.35) | 1.00 (1.00 - 1.00) | 0.492 |
| post-op TRAP test AUC (con) | 117.66 (39.25) | 107.12 (38.86) | 115.65 (39.38) | 0.99 (0.99 - 1.00) | <0.001 |
| post-op ASPI test AUC (con) | 29.32 (25.21) | 23.91 (20.93) | 28.29 (24.54) | 0.99 (0.98 - 0.99) | <0.001 |
| post-op ADP test AUC (con) | 57.92 (29.50) | 46.40 (25.24) | 55.73 (29.09) | 0.98 (0.98 - 0.99) | <0.001 |
| Haemoglobin (pre-operative) | 13.55 (1.61) | 13.27 (1.87) | 13.49 (1.66) | 0.90 (0.85 - 0.96) | 0.001 |
| pre-op Haematocrit (con) | 0.36 (0.04) | 0.35 (0.05) | 0.36 (0.04) | 0.00 (0.00 - 0.02) | <0.001 |
| pre-op mean corpuscular volume (con) | 85.62 (5.15) | 85.79 (5.48) | 85.66 (5.22) | 1.01 (0.99 - 1.03) | 0.535 |
| pre-op mean corpuscular haemoglobin (con) | 30.02 (1.90) | 30.20 (2.03) | 30.06 (1.93) | 1.05 (1.00 - 1.11) | 0.075 |
| pre-op Absolute reticulocytes (con) | 48.43 (19.61) | 47.19 (23.28) | 48.19 (20.37) | 1.00 (0.99 - 1.00) | 0.238 |
| Transferrin (con) | 1.97 (0.33) | 1.93 (0.37) | 1.96 (0.34) | 0.72 (0.52 - 0.99) | 0.046 |
| Transferrin Saturation (con) | 25.94 (11.81) | 25.91 (11.81) | 25.93 (11.81) | 1.00 (0.99 - 1.01) | 0.968 |
| Iron (con) | 12.60 (5.16) | 12.28 (5.07) | 12.54 (5.14) | 0.99 (0.97 - 1.01) | 0.272 |
| Ferritin (con) | 120.24 (112.13) | 138.13 (130.05) | 123.72 (116.02) | 1.00 (1.00 - 1.00) | 0.006 |
| pre-op lymphocyte count (con) | 1.84 (2.02) | 1.63 (1.20) | 1.80 (1.89) | 0.75 (0.64 - 0.89) | 0.001 |
| pre-op monocyte count (con) | 0.61 (0.23) | 0.60 (0.28) | 0.61 (0.24) | 0.81 (0.51 - 1.26) | 0.344 |
| pre-op neutrophil/lymphocyte Ratio (con) | 2.80 (1.97) | 3.20 (2.13) | 2.87 (2.01) | 1.09 (1.04 - 1.14) | <0.001 |
| pre-op Platelets (con) | 212.14 (62.38) | 203.27 (64.47) | 210.41 (62.88) | 1.00 (1.00 - 1.00) | 0.006 |
| pre-op IPF (con) | 3.22 (2.07) | 3.25 (2.27) | 3.23 (2.11) | 1.01 (0.96 - 1.06) | 0.740 |
| pre-op MPV (con) | 10.65 (0.99) | 10.52 (0.96) | 10.63 (0.99) | 0.87 (0.78 - 0.97) | 0.009 |
| Bilirubin (con) | 8.59 (5.52) | 9.32 (7.06) | 8.73 (5.86) | 1.02 (1.00 - 1.04) | 0.027 |
| Albumin (con) | 36.41 (3.40) | 35.68 (3.97) | 36.27 (3.53) | 0.94 (0.91 - 0.97) | <0.001 |
| Baseline eGFR (con) | 71.00 (20.35) | 69.09 (20.85) | 70.63 (20.46) | 1.00 (0.99 - 1.00) | 0.067 |
| Alkaline Phosphatase (con) | 58.81 (22.99) | 63.95 (70.16) | 59.81 (37.23) | 1.00 (1.00 - 1.01) | 0.046 |
| Troponin (con) | 9.77 (28.54) | 17.75 (42.57) | 11.97 (33.11) | 1.01 (1.00 - 1.01) | 0.110 |
| Interleukin 6 (con) | 3.04 (6.91) | 2.91 (5.12) | 3.02 (6.59) | 1.00 (0.98 - 1.01) | 0.723 |
| Interleukin 8 (con) | 3.73 (8.99) | 4.01 (7.43) | 3.79 (8.70) | 1.00 (0.99 - 1.02) | 0.567 |
| CRP (con) | 1,621,968.31 (5,029,349.96) | 1,940,144.47 (5,168,944.60) | 1,684,900.58 (5,057,479.49) | 1.00 (1.00 - 1.00) | 0.291 |
| CXCL11 (Eotaxin) | 152.89 (422.82) | 138.22 (220.56) | 150.03 (391.68) | 1.00 (1.00 - 1.00) | 0.511 |
| CX3CL1 (Fractalkine) | 70,680.92 (980,080.82) | 48,940.11 (553,009.67) | 66,448.68 (912,665.22) | 1.00 (1.00 - 1.00) | 0.680 |
| GDF-15 (mito dysfunction) | 859.12 (1,110.19) | 977.34 (1,024.68) | 882.17 (1,094.80) | 1.00 (1.00 - 1.00) | 0.069 |
| CXCL9 (iAge) | 1,376.95 (4,058.49) | 1,462.13 (2,691.89) | 1,393.54 (3,830.30) | 1.00 (1.00 - 1.00) | 0.695 |

Note: Mean (Standard deviation) for continuous variables.

**eTable 3.** Predictive accuracy of TEG for the corresponding Post-Protamine laboratory tests of coagulation/multiplate aggregometry/Sysmex parameters.

| TEG parameters (index test) | Test parameters | AUROC (Index test as binary variable) | AUROC (Index test as continuous variable) |
| --- | --- | --- | --- |
| TEG CK rmin | post-op HB | 0.503 (0.476 - 0.529) | 0.718 (0.641 - 0.795) |
| TEG CK rmin | post-op Haematocrit | 0.511 (0.508 - 0.514) | 0.664 (0.564 - 0.764) |
| TEG CK rmin | Post-op APTT | 0.519 (0.509 - 0.529) | 0.619 (0.590 - 0.648) |
| TEG CK rmin | Post-op PT (14s) | 0.508 (0.493 - 0.523) | 0.519 (0.471 - 0.568) |
| TEG CK rmin | Post-op PT (15s) | 0.504 (0.489 - 0.519) | 0.509 (0.441 - 0.576) |
| TEG CK rmin | Endogenous thrombin potential | 0.515 (0.511 - 0.520) | 0.597 (0.570 - 0.624) |
| CKH rmin | Anti-Xa heparin | * | 0.852 (0.672 - 1.000) |
| CKH rmin | Post-op APTT | * | 0.607 (0.578 - 0.636) |
| TEG CK $\alpha$ angle | Endogenous thrombin potential | 0.502 (0.499 - 0.505) | 0.570 (0.542 - 0.599) |
| TEG CK $\alpha$ angle | Clauss Fibrinogen | 0.503 (0.500 - 0.507) | 0.635 (0.611 - 0.659) |
| TEG CK $\alpha$ angle | post-op TRAP test AUC | 0.503 (0.501 - 0.505) | 0.563 (0.505 - 0.621) |
| TEG CK $\alpha$ angle | post-op ASPI test AUC | 0.504 (0.502 - 0.507) | 0.596 (0.570 - 0.623) |
| TEG CK $\alpha$ angle | post-op ADP test AUC | 0.502 (0.497 - 0.507) | 0.576 (0.546 - 0.607) |
| TEG CK $\alpha$ angle | post-op Platelets | 0.503 (0.500 - 0.507) | 0.650 (0.627 - 0.673) |
| TEG CK $\alpha$ angle | post-op MPV | 0.506 (0.499 - 0.514) | 0.522 (0.486 - 0.559) |
| TEG CK $\alpha$ angle | post-op IPF | 0.504 (0.502 - 0.506) | 0.509 (0.482 - 0.537) |
| TEG CK $\alpha$ angle | Post-op MPV X Platelets | 0.501 (0.498 - 0.504) | 0.653 (0.629 - 0.676) |
| TEG CK $\alpha$ angle | Von Willebrand factor ristocetin cofactor | 0.503 (0.502 - 0.505) | 0.673 (0.523 - 0.822) |
| TEG CK MA | Clauss Fibrinogen | 0.511 (0.506 - 0.517) | 0.829 (0.812 - 0.847) |
| TEG CK MA | D dimer | 0.502 (0.497 - 0.507) | 0.624 (0.599 - 0.649) |
| TEG CK MA | Factor XIII | 0.538 (0.522 - 0.554) | 0.741 (0.709 - 0.772) |
| TEG CK MA | Von Willebrand factor ristocetin cofactor | 0.507 (0.504 - 0.509) | 0.711 (0.594 - 0.829) |
| TEG CK MA | post-op Platelets | 0.511 (0.506 - 0.515) | 0.795 (0.776 - 0.813) |
| TEG CK MA | post-op MPV | 0.502 (0.494 - 0.510) | 0.546 (0.510 - 0.582) |
| TEG CK MA | post-op IPF | 0.502 (0.497 - 0.507) | 0.515 (0.488 - 0.543) |
| TEG CK MA | Post-op MPV X Platelets | 0.514 (0.508 - 0.520) | 0.808 (0.789 - 0.827) |
| TEG CK MA | post-op TRAP test AUC | 0.510 (0.492 - 0.528) | 0.651 (0.597 - 0.705) |
| TEG CK MA | post-op ASPI test AUC | 0.510 (0.506 - 0.513) | 0.633 (0.607 - 0.658) |
| TEG CK MA | post-op ADP test AUC | 0.516 (0.506 - 0.525) | 0.680 (0.651 - 0.708) |

Note: \* No established cut-off threshold is available.

**eTable 4.** Predictive accuracy of ROTEM for the corresponding Post-Protamine laboratory tests of coagulation/multiplate aggregometry/Sysmex parameters.

| ROTEM parameters (index test) | Test parameters | AUROC (Index test as binary variable) | AUROC (Index test as continuous variable) |
| --- | --- | --- | --- |
| ROTEM intem CT | post-op HB | 0.508 (0.505 - 0.511) | 0.564 (0.475 - 0.652) |
| ROTEM intem CT | post-op Haematocrit | 0.508 (0.505 - 0.510) | 0.540 (0.426 - 0.654) |
| ROTEM intem CT | Post-op APTT | 0.514 (0.506 - 0.522) | 0.679 (0.652 - 0.706) |
| ROTEM intem CT | Endogenous thrombin potential | 0.507 (0.502 - 0.512) | 0.535 (0.506 - 0.563) |
| ROTEM extem CT | post-op HB | 0.506 (0.469 - 0.543) | 0.555 (0.468 - 0.642) |
| ROTEM extem CT | post-op Haematocrit | 0.523 (0.464 - 0.582) | 0.617 (0.526 - 0.707) |
| ROTEM extem CT | Post-op PT (14s) | 0.558 (0.530 - 0.585) | 0.743 (0.702 - 0.783) |
| ROTEM extem CT | Post-op PT (15s) | 0.563 (0.518 - 0.608) | 0.788 (0.731 - 0.845) |
| ROTEM extem CT | Endogenous thrombin potential | 0.509 (0.500 - 0.518) | 0.521 (0.494 - 0.549) |
| ROTEM heptem CT | Anti-Xa heparin | * | 0.765 (0.609 - 0.921) |
| ROTEM heptem CT | Post-op APTT | * | 0.675 (0.648 - 0.702) |
| ROTEM intem α angle | Endogenous thrombin potential | 0.515 (0.496 - 0.535) | 0.535 (0.506 - 0.564) |
| ROTEM intem α angle | Clauss Fibrinogen | 0.666 (0.649 - 0.682) | 0.848 (0.831 - 0.864) |
| ROTEM intem α angle | post-op Platelets | 0.667 (0.651 - 0.682) | 0.848 (0.832 - 0.864) |
| ROTEM intem α angle | post-op MPV | 0.505 (0.481 - 0.529) | 0.515 (0.479 - 0.552) |
| ROTEM intem α angle | post-op IPF | 0.526 (0.506 - 0.546) | 0.562 (0.535 - 0.589) |
| ROTEM intem α angle | Post-op MPV X Platelets | 0.703 (0.684 - 0.721) | 0.838 (0.821 - 0.855) |
| ROTEM intem α angle | post-op TRAP test AUC | 0.599 (0.548 - 0.650) | 0.638 (0.580 - 0.697) |
| ROTEM intem α angle | post-op ASPI test AUC | 0.563 (0.547 - 0.579) | 0.613 (0.588 - 0.639) |
| ROTEM intem α angle | post-op ADP test AUC | 0.607 (0.582 - 0.632) | 0.657 (0.627 - 0.687) |
| ROTEM fibtem MCF | Clauss Fibrinogen | 0.617 (0.603 - 0.631) | 0.908 (0.895 - 0.920) |
| ROTEM intem MCF | Clauss Fibrinogen | 0.538 (0.530 - 0.547) | 0.871 (0.856 - 0.886) |
| ROTEM intem MCF | D dimer | 0.502 (0.493 - 0.510) | 0.618 (0.592 - 0.643) |
| ROTEM intem MCF | Factor XIII | 0.585 (0.562 - 0.608) | 0.758 (0.727 - 0.788) |
| ROTEM intem MCF | Von Willebrand factor ristocetin cofactor | 0.519 (0.515 - 0.523) | 0.625 (0.491 - 0.760) |
| ROTEM intem MCF | post-op Platelets | 0.536 (0.528 - 0.544) | 0.848 (0.832 - 0.864) |
| ROTEM intem MCF | post-op MPV | 0.503 (0.491 - 0.515) | 0.529 (0.492 - 0.565) |
| ROTEM intem MCF | post-op IPF | 0.504 (0.495 - 0.514) | 0.512 (0.484 - 0.539) |
| ROTEM intem MCF | Post-op MPV X Platelets | 0.546 (0.536 - 0.557) | 0.859 (0.843 - 0.875) |
| ROTEM intem MCF | post-op TRAP test AUC | 0.537 (0.505 - 0.570) | 0.672 (0.617 - 0.727) |
| ROTEM intem MCF | post-op ASPI test AUC | 0.522 (0.515 - 0.528) | 0.632 (0.606 - 0.657) |
| ROTEM intem MCF | post-op ADP test AUC | 0.544 (0.529 - 0.560) | 0.675 (0.646 - 0.704) |

Note: \* No established cut-off threshold is available.

**eTable 5.**
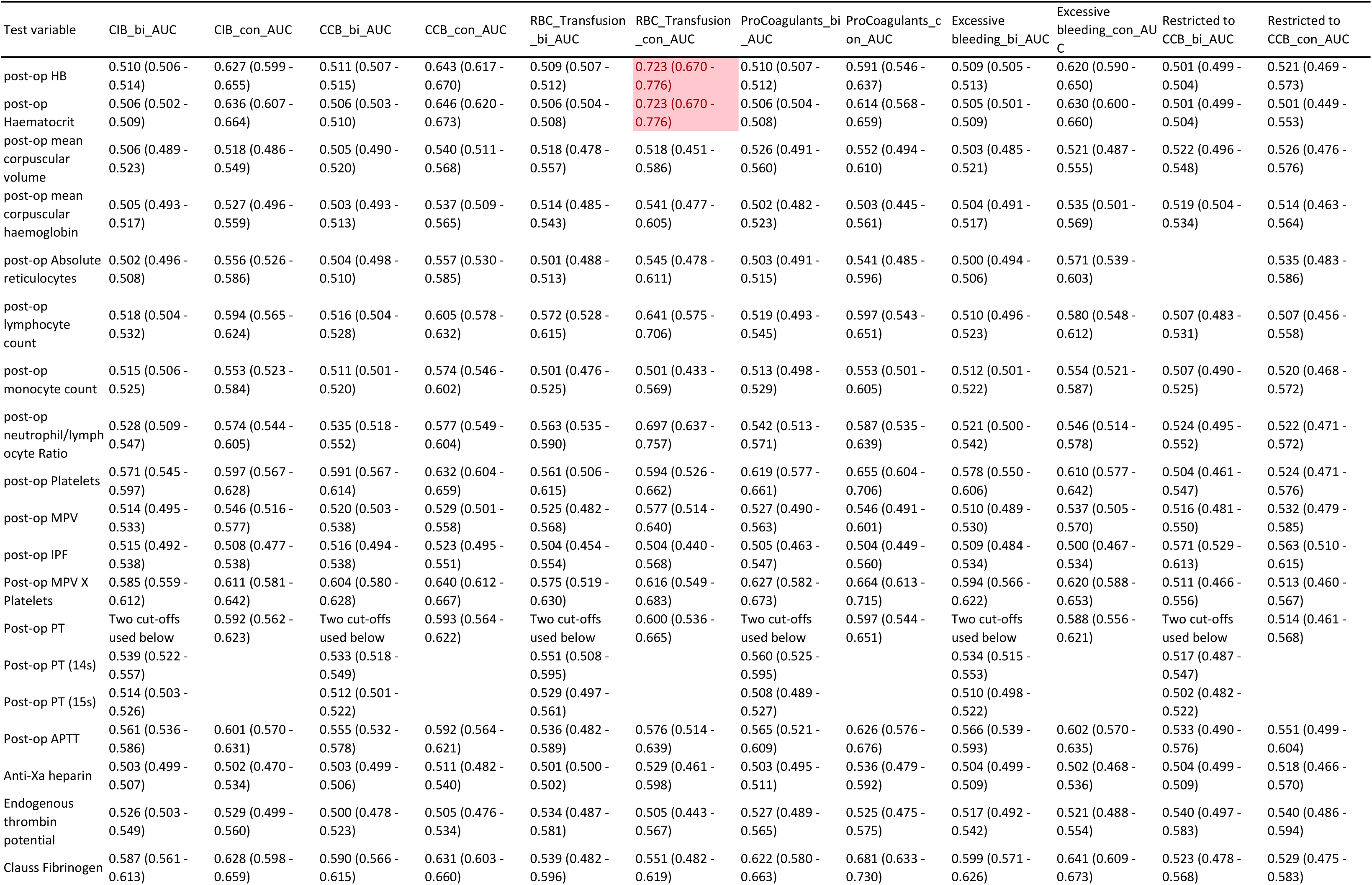
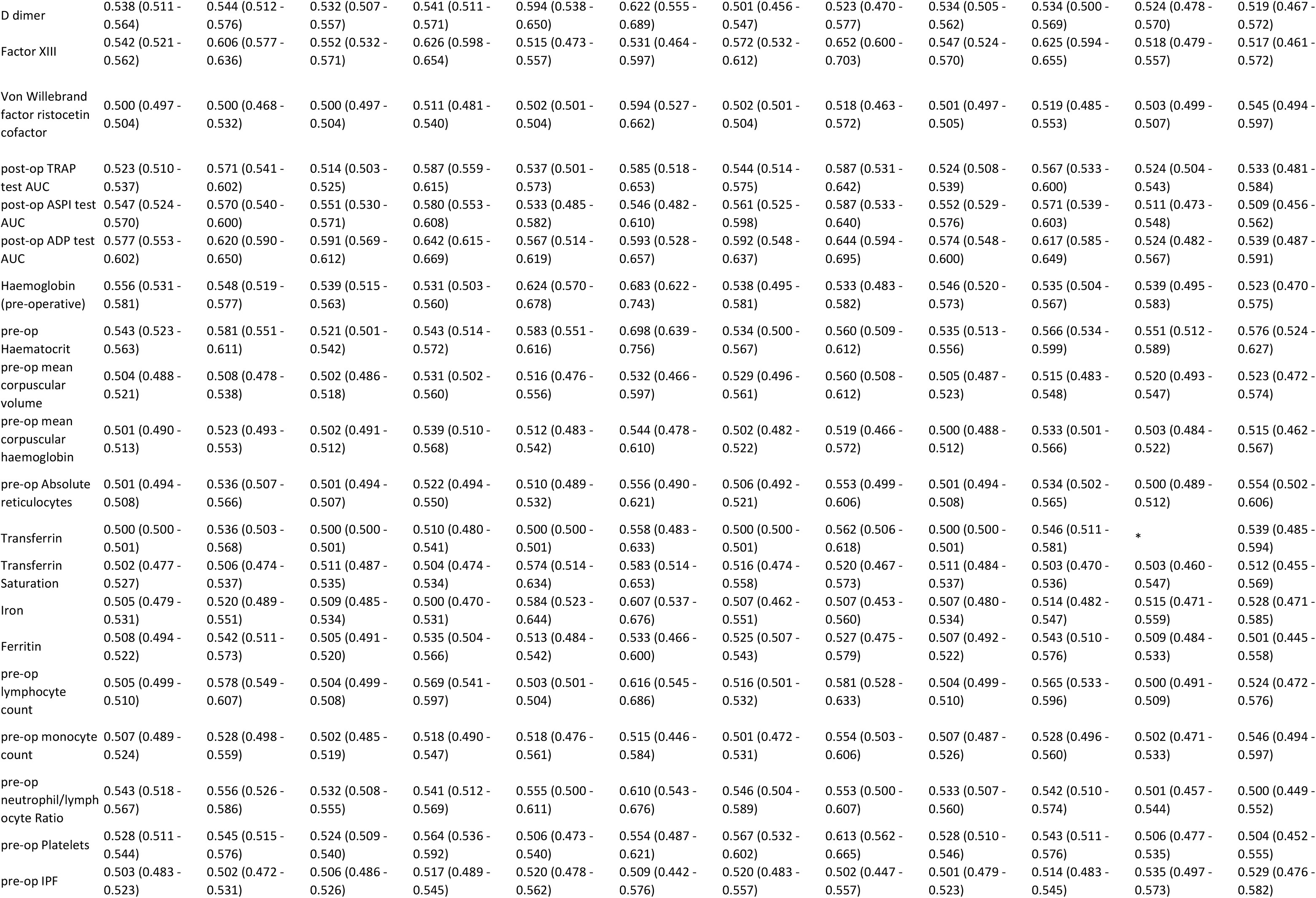
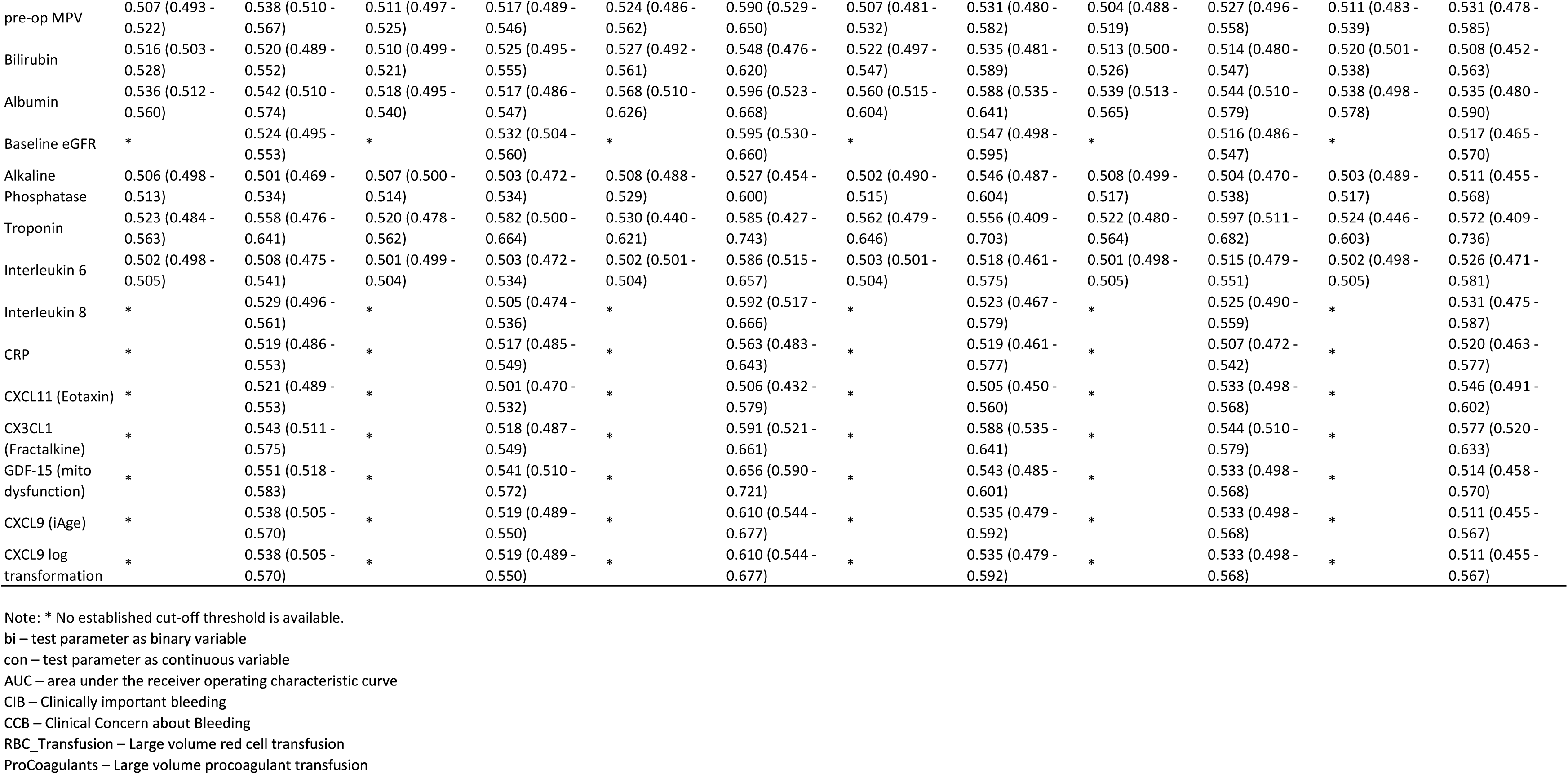
Diagnostic accuracy of individual tests (continuous and binary) for clinically important bleeding, its three individual components, and the secondary outcome. Note: the tests include laboratory tests of coagulation, Sysmex full blood count, multiplate aggregometry, baseline biomarkers of multimorbidity and biological ageing

**eTable 6.** Predictors, discrimination (AUROC and 95% CI), and calibration (Brier score and O:E ratio) for models to predict clinically important bleeding and its three individual components.

| Outcome | Model name | Predictors | AUC (95% CI) | Brier score | O:E ratio |
| --- | --- | --- | --- | --- | --- |
| Clinically important bleeding | TEG | TEG CK MA, TEG CK $\alpha$ angle, | 0.611 (0.521 - 0.701) | 0.154 | 1.003 |
| Clinically important bleeding | ROTEM | ROTEM intem $\alpha$ angle, | 0.588 (0.496 - 0.681) | 0.154 | 1.002 |
| Clinically important bleeding | Organ dysfunction biomarkers | Albumin, Haematocrit (Preop), absolute neutrophils (Preop), IPF (Preop), MPV (Preop), mean corpuscular haemoglobin (Preop), Platelets (Preop), Ferritin, | 0.606 (0.512 - 0.701) | 0.152 | 1.004 |
| Clinically important bleeding | Ageing biomarkers | cxcl9 log2 transformation, | 0.536 (0.433 - 0.639) | 0.157 | 1.006 |
| Clinically important bleeding | Lab tests: post-op (Sysmex + Multiplate + Coagulation) | Haematocrit (Postop), APTT, Clauss Fibrinogen, AUC ADP (Postop), D dimer, neutrophil/lymphocyte Ratio (Postop), | 0.688 (0.607 - 0.769) | 0.147 | 1.006 |
| Clinically important bleeding | Organ dysfunction biomarkers + Ageing biomarkers + Lab tests | absolute neutrophils (Preop), APTT, Clauss Fibrinogen, Haematocrit (Preop), D dimer, MPV (Postop), Ferritin, IPF (Preop), AUC ADP (Postop), Albumin, | 0.701 (0.620 - 0.781) | 0.144 | 1.007 |
| Clinically important bleeding | TEG + Organ dysfunction biomarkers + Ageing biomarkers + Lab tests | Ferritin, Haematocrit (Preop), absolute neutrophils (Postop), APTT, Clauss Fibrinogen, Iron, D dimer, AUC ADP (Postop), | 0.694 (0.612 - 0.775) | 0.145 | 1.008 |
| Clinically important bleeding | ROTEM + Organ dysfunction biomarkers + Ageing biomarkers + Lab tests | Haematocrit (Preop), Clauss Fibrinogen, D dimer, Platelets (Preop), Iron, MPV (Postop), Ferritin, AUC ADP (Postop), absolute neutrophils (Preop), mean corpuscular haemoglobin (Postop), | 0.692 (0.610 - 0.775) | 0.145 | 1.008 |
| Large volume red cell transfusion | ROTEM | ROTEM fibtem MCF, ROTEM intem $\alpha$ angle, | 0.580 (0.392 - 0.766) | 0.036 | 1.030 |
| Large volume red cell transfusion | Organ dysfunction biomarkers | haemoglobin (Preop), Albumin, IPF (Preop), MPV (Preop), Platelets (Preop), Interleukin 8, | 0.701 (0.501 - 0.900) | 0.035 | 1.026 |
| Large volume red cell transfusion | Ageing biomarkers | GDF-15, cxcl9 log2 transformation, | 0.646 (0.410 - 0.880) | 0.035 | 1.043 |
| Large volume red cell transfusion | Lab tests: post-op (Sysmex + Multiplate + Coagulation) | Platelets (Postop), Haematocrit (Postop), absolute neutrophils (Postop), | 0.745 (0.602 - 0.886) | 0.034 | 1.041 |
| Large volume red cell transfusion | Organ dysfunction biomarkers + Ageing biomarkers + Lab tests | absolute neutrophils (Postop), Platelets (Postop), Albumin, MPV (Preop), IPF (Preop), Haematocrit (Postop), | 0.754 (0.602 - 0.903) | 0.034 | 1.047 |
| Large volume red cell transfusion | TEG + Organ dysfunction biomarkers + Ageing biomarkers + Lab tests | haemoglobin (Preop), Platelets (Postop), absolute neutrophils (Postop), Haematocrit (Postop), Albumin, | 0.762 (0.626 - 0.897) | 0.034 | 1.037 |
| Large volume red cell transfusion | ROTEM + Organ dysfunction biomarkers + Ageing biomarkers + Lab tests | Albumin, absolute neutrophils (Postop), Haematocrit (Postop), | 0.753 (0.608 - 0.895) | 0.034 | 1.039 |
| Large volume procoagulant transfusion | TEG | TEG CK MA, | 0.649 (0.501 - 0.798) | 0.053 | 1.008 |
| Large volume procoagulant transfusion | ROTEM | ROTEM intem MCF, | 0.630 (0.487 - 0.774) | 0.053 | 1.008 |
| Large volume procoagulant transfusion | Organ dysfunction biomarkers | mean corpuscular volume (Preop), Albumin, Platelets (Preop), Baseline eGFR, mean corpuscular haemoglobin (Preop), neutrophil/lymphocyte Ratio (Preop), | 0.651 (0.497 - 0.805) | 0.053 | 1.010 |
| Large volume procoagulant transfusion | Lab tests: post-op (Sysmex + Multiplate + Coagulation) | AUC ADP (Postop), APTT, Clauss Fibrinogen, mean corpuscular volume (Postop), D dimer, haemoglobin (Postop), Haematocrit (Postop), neutrophil/lymphocyte Ratio (Postop), | 0.720 (0.595 - 0.845) | 0.052 | 1.020 |
| Large volume procoagulant transfusion | Organ dysfunction biomarkers + Ageing biomarkers + Lab tests | Haematocrit (Postop), APTT, AUC ADP (Postop), Platelets (Postop), haemoglobin (Postop), Factor XIII, mean corpuscular volume (Postop), Albumin, absolute neutrophils (Postop), | 0.723 (0.590 - 0.857) | 0.052 | 1.025 |
| Large volume<br>procoagulant transfusion | TEG + Organ dysfunction<br>biomarkers + Ageing<br>biomarkers + Lab tests | Platelets (Preop), haemoglobin (Preop), AUC ADP (Postop),<br>Albumin, absolute neutrophils (Postop), APTT, Clauss<br>Fibrinogen, Haematocrit (Postop), haemoglobin (Postop), | 0.732 (0.606 - 0.858) | 0.051 | 1.021 |
| Large volume<br>procoagulant transfusion | ROTEM + Organ dysfunction<br>biomarkers + Ageing<br>biomarkers + Lab tests | Platelets (Preop), APTT, Clauss Fibrinogen, absolute<br>neutrophils (Postop), Haematocrit (Postop), haemoglobin<br>(Postop), AUC ADP (Postop), haemoglobin (Preop),<br>Albumin, | 0.732 (0.606 - 0.858) | 0.051 | 1.021 |
| Severe blood loss | TEG | TEG CK MA, TEG CK $\alpha$ angle, | 0.620 (0.523 - 0.716) | 0.134 | 1.005 |
| Severe blood loss | ROTEM | ROTEM intem MCF, | 0.606 (0.508 - 0.704) | 0.135 | 1.003 |
| Severe blood loss | Organ dysfunction biomarkers | Albumin, Haematocrit (Preop), absolute neutrophils<br>(Preop), MPV (Preop), Bilirubin, Alkaline Phosphatase, IPF<br>(Preop), mean corpuscular haemoglobin (Preop), Absolute<br>reticulocytes (Preop), Platelets (Preop), | 0.602 (0.501 - 0.702) | 0.134 | 1.006 |
| Severe blood loss | Ageing biomarkers | cxcl9 log2 transformation, | 0.530 (0.419 - 0.641) | 0.138 | 0.999 |
| Severe blood loss | Lab tests: post-op (Sysmex +<br>Multiplate + Coagulation) | AUC ADP (Postop), APTT, Clauss Fibrinogen, D dimer,<br>haemoglobin (Postop), mean corpuscular haemoglobin<br>(Postop), | 0.680 (0.592 - 0.769) | 0.130 | 1.007 |
| Severe blood loss | Organ dysfunction biomarkers<br>+ Ageing biomarkers + Lab<br>tests | Absolute reticulocytes (Postop), APTT, Clauss Fibrinogen,<br>MPV X Platelets (Postop), Haematocrit (Preop), Ferritin,<br>Albumin, IPF (Preop), AUC ADP (Postop), Platelets<br>(Postop), absolute neutrophils (Preop), | 0.701 (0.613 - 0.788) | 0.127 | 1.007 |
| Severe blood loss | TEG + Organ dysfunction<br>biomarkers + Ageing<br>biomarkers + Lab tests | AUC ADP (Postop), Absolute reticulocytes (Postop),<br>Haematocrit (Preop), Ferritin, APTT, Clauss Fibrinogen,<br>Iron, D dimer, | 0.686 (0.596 - 0.776) | 0.128 | 1.008 |
| Severe blood loss | ROTEM + Organ dysfunction<br>biomarkers + Ageing<br>biomarkers + Lab tests | Iron, absolute neutrophils (Preop), Clauss Fibrinogen, D<br>dimer, Haematocrit (Preop), Ferritin, Absolute<br>reticulocytes (Postop), mean corpuscular haemoglobin<br>(Postop), AUC ADP (Postop), Albumin, | 0.695 (0.606 - 0.783) | 0.127 | 1.011 |

## Footnotes

1 QUADAS-2 http://www.bristol.ac.uk/media-library/sites/quadas/migrated/documents/quadas2.pdf

2 https://www.isrctn.com/ISRCTN20630689

## References

1. Ranucci M, Bozzetti G, Ditta A, Cotza M, Carboni G, Ballotta A, (2008) Surgical reexploration after cardiac operations: why a worse outcome? Ann Thorac Surg 86: 1557–1562

2. Karkouti K, Wijeysundera DN, Yau TM, Beattie WS, Abdelnaem E, McCluskey SA, Ghannam M, Yeo E, Djaiani G, Karski J, (2004) The independent association of massive blood loss with mortality in cardiac surgery. Transfusion 44: 1453–1462

3. Serraino GF, Murphy GJ, (2017) Routine use of viscoelastic blood tests for diagnosis and treatment of coagulopathic bleeding in cardiac surgery: updated systematic review and meta-analysis. Br J Anaesth 118: 823–833

4. Roman MA, Abbasciano RG, Pathak S, Oo S, Yusoff S, Wozniak M, Qureshi S, Lai FY, Kumar T, Richards T, Yao G, Estcourt L, Murphy GJ, (2021) Patient blood management interventions do not lead to important clinical benefits or cost-effectiveness for major surgery: a network meta-analysis. Br J Anaesth 126: 149–156

5. Dyke C, Aronson S, Dietrich W, Hofmann A, Karkouti K, Levi M, Murphy GJ, Sellke FW, Shore-Lesserson L, von Heymann C, Ranucci M, (2014) Universal definition of perioperative bleeding in adult cardiac surgery. J Thorac Cardiovasc Surg 147: 1458–1463 e1451

6. Murphy GJ, Mumford AD, Rogers CA, Wordsworth S, Stokes EA, Verheyden V, Kumar T, Harris J, Clayton G, Ellis L, Plummer Z, Dott W, Serraino F, Wozniak M, Morris T, Nath M, Sterne JA, Angelini GD, Reeves BC, (2017) Diagnostic and therapeutic medical devices for safer blood management in cardiac surgery: systematic reviews, observational studies and randomised controlled trials. Programme Grants Appl Res 5

7. Wozniak MJ, Abbasciano R, Monaghan A, Lai FY, Corazzari C, Tutino C, Kumar T, Whiting P, Murphy GJ, (2021) Systematic Review and Meta-Analysis of Diagnostic Test Accuracy Studies Evaluating Point-of-Care Tests of Coagulopathy in Cardiac Surgery. Transfus Med Rev 35: 7–15

8. Lai FY, Adebayo AS, Sheikh S, Roman M, Joel-David L, Aujla H, Chad T, Tomkova K, Ladak S, Condorelli G, Zakkar M, Solomon C, Woźniak MJ, Murphy GJ, (2024) Immune system homeostasis in people with multiple long-term conditions determines susceptibility to organ injury and mortality following cardiac surgery. medRxiv

9. Bossuyt PM, Reitsma JB, Bruns DE, Gatsonis CA, Glasziou PP, Irwig L, Lijmer JG, Moher D, Rennie D, de Vet HC, Kressel HY, Rifai N, Golub RM, Altman DG, Hooft L, Korevaar DA, Cohen JF, Group S, (2015) STARD 2015: an updated list of essential items for reporting diagnostic accuracy studies. BMJ 351: h5527

10. Collins GS, Reitsma JB, Altman DG, Moons KG, (2015) Transparent Reporting of a multivariable prediction model for Individual Prognosis or Diagnosis (TRIPOD): the TRIPOD statement. Ann Intern Med 162: 55–63

11. Rubin D (2004) Multiple imputation for nonresponse in surveys. Wiley, Canada

12. Vickers AJ, Van Calster B, Steyerberg EW, (2016) Net benefit approaches to the evaluation of prediction models, molecular markers, and diagnostic tests. BMJ 352: i6

13. Mandrekar JN, (2010) Receiver operating characteristic curve in diagnostic test assessment. J Thorac Oncol 5: 1315–1316

14. Fischer JE, Bachmann LM, Jaeschke R, (2003) A readers’ guide to the interpretation of diagnostic test properties: clinical example of sepsis. Intensive Care Med 29: 1043–1051

15. Gurbel PA, Bliden KP, Tantry US, Monroe AL, Muresan AA, Brunner NE, Lopez-Espina CG, Delmenico PR, Cohen E, Raviv G, Haugen DL, Ereth MH, (2016) First report of the point-of-care TEG: A technical validation study of the TEG-6S system. Platelets 27: 642–649

16. Neal MD, Moore EE, Walsh M, Thomas S, Callcut RA, Kornblith LZ, Schreiber M, Ekeh AP, Singer AJ, Lottenberg L, Foreman M, Evans S, Winfield RD, Goodman MD, Freeman C, Milia D, Saillant N, Hartmann J, Achneck HE, (2020) A comparison between the TEG 6s and TEG 5000 analyzers to assess coagulation in trauma patients. J Trauma Acute Care Surg 88: 279–285

17. Qureshi SH, Patel NN, Murphy GJ, (2018) Vascular endothelial cell changes in postcardiac surgery acute kidney injury. Am J Physiol Renal Physiol 314: F726–F735

