## Supplementary material for "Predictive accuracy of diagnostic tests for excessive bleeding in cardiac surgery: the COPTIC-C study": COPTIC-C. Supplementary materials

[1] Robert Grant (Clinical Research Fellow)

[1] Florence Y Lai (Senior Statistician)

[1] Hardeep Aujla (Senior Research Manager)

[1] Marcin Wozniak (Lecturer in Cardiovascular Sciences)

[1] Hasmukh R Patel (Senior Laboratory Technician)

[2,3] Laura Green (Professor of Haemostasis and Transfusion Medicine) ORCID: 0000-0003-4063-9768

[4] Andrew Mumford (Professor of Haematology)

[1] Gavin J Murphy (British Heart Foundation Chair of Cardiac Surgery, Chief Investigator)

**Affiliations**

[1] Department of Cardiovascular Sciences, University of Leicester, UK

[2] Blizard Institute, Queen Mary University of London, UK

[3] NHS Blood and Transplant, London, UK

[4] Bristol Heart Institute, University of Bristol, UK

**Corresponding Author**

Dr Weiqi Liao, Senior Statistician, Department of Cardiovascular Sciences, University of Leicester, Clinical Sciences Wing, Glenfield Hospital, Leicester LE3 9QP.

### Tables for the Methods section

#### eTable A – The thresholds for Pre-Surgery and Post-Protamine Sysmex testing and Multiplate Aggregometry, and Post-Protamine Laboratory Tests for Coagulopathy

| Test | Assay | Units | Interpretation of an abnormal result | Thresholds |
|---|---|---|---|---|
| **Sysmex tests** | | | | |
| Haemoglobin (hb) | Sysmex XE-2100 | g/dL | Low test result | Male <13.7<br>Female <12.0 |
| Haematocrit (HCt) | Sysmex XE-2100 | % | Low test result | Male <0.40<br>Female <0.37 |
| Absolute neutrophils | FBC (XE-2100 cell counter) | $X10^9/L$ | Low test result | <1 |
| Lymphocyte count | FBC (XE-2100 cell counter) | $X10^9/L$ | Low test result | <0.5 |
| Monocyte count | FBC (XE-2100 cell counter) | $X10^9/L$ | High test result | >0.8 |
| Neutrophil/Lymphocyte Ratio | Sysmex XE-2100 (Milton Keynes, UK) | | High test result | >3 |
| Mean corpuscular volume | FBC (XE-2100 cell counter) | fl | Low test result | <80 |
| Mean corpuscular haemoglobin | FBC (XE-2100 cell counter) | pg | Low test result | <27 |
| Post-op Absolute reticulocytes | Sysmex XE-2100 | % | High test result | <0.68 |
| Post-op PLT | Sysmex XE-2100 | $X10^9/L$ | Low test result | <140 |
| Post-op MPV | Sysmex XE-2100 | fL | Low test result | <9.32 |
| Post-op IPF | Sysmex XE-2100 | % | High test result | >4.32 |
| Post-op MPV x Platelets | Sysmex XE-2100 | $fL.X10^9/L$ | Low test result | <140 |
| **Multiplate tests** | | | | |
| AUC TRAP-test | Multiplate | Aggregation units (U) | Low test result | <50 |
| AUC ASPI-test | Multiplate | Aggregation units (U) | Low test result | <30 |
| AUC ADP-test | Multiplate | Aggregation units (U) | Low test result | <30 |

| Post Protamine Laboratory Tests of Coagulopathy | | | | |
|---|---|---|---|---|
| Post-op Prothrombin Time (PT) | Innovin | Seconds (s) | High test result | >15 |
| Post-op Activated Partial Thromboplastin Time (APTT) | actin FS | Seconds (s) | High test result | >35 |
| Claus Fibrinogen (Factor I) | Clauss Fibrinogen activity (thrombin reagent) | g/L | Low test result | <2 |
| anti-Xa | Hyphen Biomed anti-Xa kit | IU/mL | High test result | >1.0 |
| DDIC | Innovance D-dimer assay | ng/mL | High test result | >500 |
| FXIII | FXIII activity (Berichrom XIII), | IU/dL | Low test result | <60 |
| vWF | vWF Risctocetin cofactor activity (BC RiCof assay) | IU/dL | Low test result | <50 |
| ETP (Factor II) | FluCa-kit (Stago, Asnieres sur Seine, France) | nM/min | Low test result | <1336 |

Abbreviation

- AUC = area under the curve

### eTable B – Post-Protamine TEG criteria for a positive test result

| Parameters | Units | Interpretation of an abnormal result | Thresholds |
|---|---|---|---|
| TEG CK R | Mm | High test result | > 10 |
| TEG CK α angle | Degrees | Low test result | < 37.6 |
| TEG CK MA | Mm | Low test result | < 44 |
| TEG CKH R | Minutes (mm) | High test result | N/A |

Abbreviations

- α-Angle: measures the speed of fibrin build-up
- CK: citrated kaolin
- CKH: citrated kaolin with heparinise
- MA: maximum amplitude of the fibrin clot
- R (min): reaction time (in minutes) to initial fibrin formation

### eTable C – Post-Protamine ROTEM criteria for a positive test result

| Parameter | Units | Interpretation of an abnormal result | Thresholds |
|---|---|---|---|
| **INTEM** | | | |
| intem CT | Minutes (min) | High test result | >240 |
| intem α angle | Degrees | Low test result | <70 |
| intem MCF | Mm | Low test result | <50 |
| **EXTEM** | | | |
| extem CT | Minutes (m) | High test result | >79 |
| **FIBTEM** | | | |
| fibtem MCF | Mm | Low test result | <9 |
| **HEPTEM** | | | |
| heptem CT | Minutes (min) | High test result | N/A |

Abbreviations

- CT: clotting time
- MCF: maximum clot firmness

### eTable D – Baseline biomarkers of Organ Dysfunction and Biological Ageing

| Biomarkers | Units | Interpretation of an abnormal result | Thresholds |
|---|---|---|---|
| **Haematopoiesis** | | | |
| Baseline Haemoglobin | g/dL | Low test result | Male <13.7<br>Female <12.0 |
| Baseline haematocrit | % | Low test result | Male <0.40<br>Female< 0.37 |
| Baseline Absolute Reticulocyte | % | High test result | <0.68 |
| Lymphocyte count | $X10^9/L$ | Low test result | <0.2 |
| Baseline monocyte count | $X10^9/L$ | High test result | >0.8 |
| Baseline neutrophil/lymphocyte ratio | | High test result | >3 |
| Transferrin | g/L | | |
| Iron | µmol/L | | <10 |
| Transferrin Saturation | % | Low test result | <20% |
| Serum Ferritin | ng/mL | | Adult male <25<br>Adult female <10 |
| Baseline Platelet count | $X10^9/L$ | Low test result | <140 |
| Post-op MPV | fL | Low test result | <9.32 |
| Post-op IPF | % | High test result | >4.32 |
| Post-op MPV x Platelets | fL | Low test result | <140 |
| **Liver** | | | |
| Bilirubin | µmol/L | High test result | >20 |
| Albumin | g/L | Low test result | <34 |
| **Bone** | | | |
| Alkaline Phosphatase | IU/L | High test result | >147 |
| **Kidney** | | | |
| eGFR | mls/min×1.73m$^2$ | Low test result | <60 |
| **Inflammation** | | | |
| CRP | pg/mL | High test result | >5 |
| Interleukin 6 | pg/mL | High test result | >43.5 |

| | | |
|---|---|---|
| Interleukin 8 | pg/mL | N/A |
| **Biological Ageing** | | |
| CCL11/Eotaxin | pg/mL | N/A |
| CX3CL1/Fractalkine | pg/mL | N/A |
| GDF-15/Mito dysfunction | pg/mL | N/A |
| CXCL9/iAge | pg/mL | N/A |
